# A Qualitative Study of People with Alzheimer’s Disease in a Memory Clinic Considering Lecanemab Treatment

**DOI:** 10.1101/2024.09.17.24313315

**Authors:** Anna L Parks, Ayush Thacker, Daniel Dohan, Liliana A Ramirez Gomez, Christine S Ritchie, Joanna Paladino, Sachin J Shah

## Abstract

**Background:** People with Alzheimer’s disease (AD) now have access to disease-modifying treatment with anti-amyloid monoclonal antibodies (mAbs). Their perception of risks and benefits and approach to treatment decisions remain unknown.

**Objective:** We aimed to understand how people with AD weigh the benefits and costs of anti-amyloid mAbs and incorporate these into decisions about treatment.

**Methods:** We conducted semi-structured interviews with people with biomarker- or imaging-confirmed AD and mild or moderate cognitive impairment who were seen at memory care clinics and discussed lecanemab with a clinician. Interviews were recorded, transcribed, and deidentified. Thematic analysis identified themes and subthemes.

**Results:** Among 22 participants (mean age 70, 8 [36%] women, 22 [100%] White), analysis revealed 3 major themes and associated subthemes: 1) People with AD sought and obtained information from different sources—advocacy organizations, the Internet, and clinicians; 2) Hopes, expected benefits, and the existential threat of dementia drove willingness and readiness to start lecanemab; 3) Individual traits, family factors, and degree of trust in expertise influenced how people balanced risks and benefits. Some would accept treatment at any cost; others carefully weighed risks and burdens, but were motivated by supportive families, insurance coverage, and trust in expertise; for a few, costs decidedly outweighed their personal benefits. People with AD desired more individualized information and to hear more from patients who took the medication.

**Conclusion:** Results from this first qualitative study of people with AD considering treatment with anti-amyloid mAbs can inform clinician, health system and policy efforts to individualize decisions.

## Introduction

Approval of anti-amyloid monoclonal antibodies (mAbs) is a major advance in the field of Alzheimer’s disease (AD). This medication class is the first potentially disease-modifying therapy for people with AD. Lecanemab, approved by the Food and Drug Administration (FDA) in 2023, can slow cognitive decline in early AD by six months.^1^ Donanemab was FDA-approved in July 2024, and clinical trials of additional mAbs are in process.^2^

Alongside therapeutic optimism is growing recognition of potential tradeoffs that complicate treatment decisions. Side effects include intracranial swelling (amyloid-related imaging abnormalities-edema), and intracranial bleeding (amyloid-related imaging abnormalities-hemorrhage [ARIA-H]).^3^ Certain comorbidities and medications, like anticoagulants, and genotypes, like apolipoprotein E ε4 (*ApoE4*), increase the ARIA-H risk.^4^ Although most ARIA is asymptomatic, it can result in death and disability.^5^ How patients fare long-term remains unknown. Treatment requires time expenditure, including subspecialty visits, diagnostic testing, monitoring, and infusions every two-to-four weeks. And anti-amyloid mAbs are financially costly for individuals and society.^6^

Many have called for shared decision-making to address the complexity of treatment decisions.^4,5,7–9^ To be effective, shared decision-making requires understanding how patients perceive the risks and benefits of treatment and how they approach decisions.^10^ In the new era of anti-amyloid mAbs, our aim was to understand how people with AD weigh potential benefits of treatment against costs and how they make treatment decisions.

## Methods

### Study Design and Data Collection

We conducted an exploratory qualitative study using semi-structured interviews of people seen in memory clinics with AD between February-March 2024. A multidisciplinary team developed an interview guide using the Ottawa Decision Support Framework.^11^ The guide included questions about 1) information received, 2) hopes, motivations, and concerns about treatment, 3) level of certainty about starting treatment, 4) factors influencing decision-making, and 5) the decision-making process. Interviews were conducted remotely by Zoom or telephone by trained study personnel (A.P., A.T.). The institutional review boards of participating institutions approved this study as exempt. We adhered to Consolidated Criteria for Reporting Qualitative Studies (COREQ) guidance.^12^

### Setting and Participants

Participants were recruited by mail and telephone from memory clinic referral centers at two academic medical centers, the University of Utah and Massachusetts General Hospital. **Figure 1** shows a flowchart of recruitment. We screened memory clinic referrals and included subjects who:1) were English-speakers, 2) between ages 50-90 years, 3) had at least one memory clinic visit where lecanemab was discussed, 4) underwent initial diagnostic testing to confirm AD and assess for lecanemab eligibility (e.g., amyloid on cerebrospinal fluid and/or positron-emission tomography, magnetic resonance imaging and *ApoE4 testing*). To reflect the full breadth of patients with whom clinicians have shared decision-making discussions, neither consent for nor subsequent receipt of lecanemab were inclusion criteria. We included subjects with Montreal Cognitive Assessment (MOCA) scores ≥16 (mild or moderate cognitive impairment), consistent with current appropriate use criteria for lecanemab.^5^ We included two participants with biomarker or imaging-confirmed AD who had normal MOCA score (≥26) but clinical diagnosis of mild cognitive impairment based on Quick Dementia Rating System or Clinical Dementia Rating-Sum of Boxes score. These two participants were treated with lecanemab and were included to accurately represent real-world clinical practice. Participants completed verbal informed consent and an online demographic questionnaire before participation and received a $50 gift card.

**Figure 1.** Timeline of Recruitment and Interviews in Relation to Clinical Evaluation for People with AD Considering Anti-Amyloid Monoclonal Antibody Treatment.

### Data Analysis

Interviews were digitally recorded, transcribed, and de-identified. Qualitative thematic analysis was performed in March-April 2024 using a stepwise approach and hybrid deductive-inductive coding methods.^13,14^ The primary analytic team was comprised of AP (female physician-researcher with qualitative research expertise), AT (male clinical research coordinator), and JP (female researcher with serious illness communication and qualitative research expertise).

Coders independently coded the first five transcripts deductively using the interview guide as a framework; additional codes were added inductively to create the preliminary codebook. AT, AP, and JP coded the remaining transcripts and met every five interviews to discuss discrepancies, resolve differences by consensus, revise codes, ensure iterative content building, and examine and address how personal experiences or perspectives might influence the analysis. The final codebook was applied to all interviews. Using meetings with two additional experts (DD, SS), codes were grouped to develop preliminary themes and sub-themes. Associations were confirmed by the entire group to identify final themes. AP, AT, and JP reviewed transcripts iteratively to assess data saturation, or when interviews were not gleaning new information, at which point recruitment was stopped.^15–17^

## Results

Characteristics for the 22 participants are provided in **Table 1**. The study cohort included 12 (55%) people from Massachusetts General Hospital and 10 (45%) from the University of Utah. Mean age was 70 years, 8 (36%) were women, and 22 (100%) were non-Hispanic White. Eight (36%) had less than a college degree, 8 (50%) had annual income less than $100,000 (6 [27%] were unsure or had no income), and 6 (27%) lived rurally. Mean Montreal Cognitive Assessment (MOCA) was 20 (standard deviation 3); 2 (9%) had normal cognitive performance (MOCA ≥26), 15 (68%) had mild cognitive impairment (MOCA 18-25), and 5 (23%) had moderate cognitive impairment (MOCA 10-17). A majority (12 [55%]) were ready to decide on mAb treatment, and the remainder were evenly split between ready but wanting more information (5 [23%]) and not ready (5 [23%]). A small proportion would decline (3 [14%]), a similar proportion needed more information (4 [18%]), and most would pursue treatment (15 [68%]).

**Table 1.** Patient Characteristics.

| Characteristic | N=22 |
| --- | --- |
| Age mean (SD) | 70 (10) |
| Institution n (%) |  |
| Massachusetts General Hospital | 12 (55%) |
| University of Utah | 10 (45%) |
| Gender n (%) |  |
| Male | 14 (64%) |
| Female | 8 (36%) |
| Orientation n (%) |  |
| Heterosexual or straight | 22 (100%) |
| Race n (%) |  |
| White | 22 (100%) |
| Ethnicity n (%) |  |
| Non-Hispanic | 22 (100%) |
| Geographic Location n (%) |  |
| Suburban | 14 (64%) |
| Rural | 6 (27%) |
| Urban | 2 (9.1%) |
| Religious Affiliation n (%) |  |
| Christian | 16 (73%) |
| Agnostic | 3 (14%) |
| Atheist | 1 (4.5%) |
| Buddhist | 1 (4.5%) |
| Prefer not to answer | 1 (4.5%) |
| Highest level of education obtained n (%) |  |
| High School | 1 (4.5%) |
| Some college | 5 (23%) |
| Associate's Degree | 2 (9.1%) |
| Bachelor's Degree | 7 (32%) |
| Master's Degree | 4 (18%) |
| Doctorate Degree | 3 (14%) |
| Household Income n (%) |  |
| \$30,000-59,999 | 1 (4.5%) |
| \$60,000-99,999 | 7 (32%) |
| \$100,000 or more | 8 (36%) |
| Unsure/No Income | 6 (27%) |
| Montreal Cognitive Assessment (MOCA) score mean (SD) | 20 (2) |
| Normal cognitive performance (MOCA $\geq 26$) n (%)* | 2 (9) |
| Mild cognitive impairment (MOCA 18-25) n (%) | 15 (68) |
| Moderate cognitive impairment (MOCA 10-17) n (%)** | 5 (23) |
| How ready do you feel to decide about treatment n (%) |  |
| Ready to make a decision | 12 (55%) |
| Ready to make a decision but has more questions | 5 (23%) |
| Not ready to make a decision | 5 (23%) |
| If you had to decide today about treatment n (%) |  |
| Yes to treatment | 15 (68%) |
| Not sure, need more information | 4 (18%) |
| No to treatment | 3 (14%) |

Through analysis, we identified three primary themes and associated subthemes (**Figure 2**), with representative quotations in the manuscript text. **Table 2** provides additional supportive quotations.

**Figure 2.**
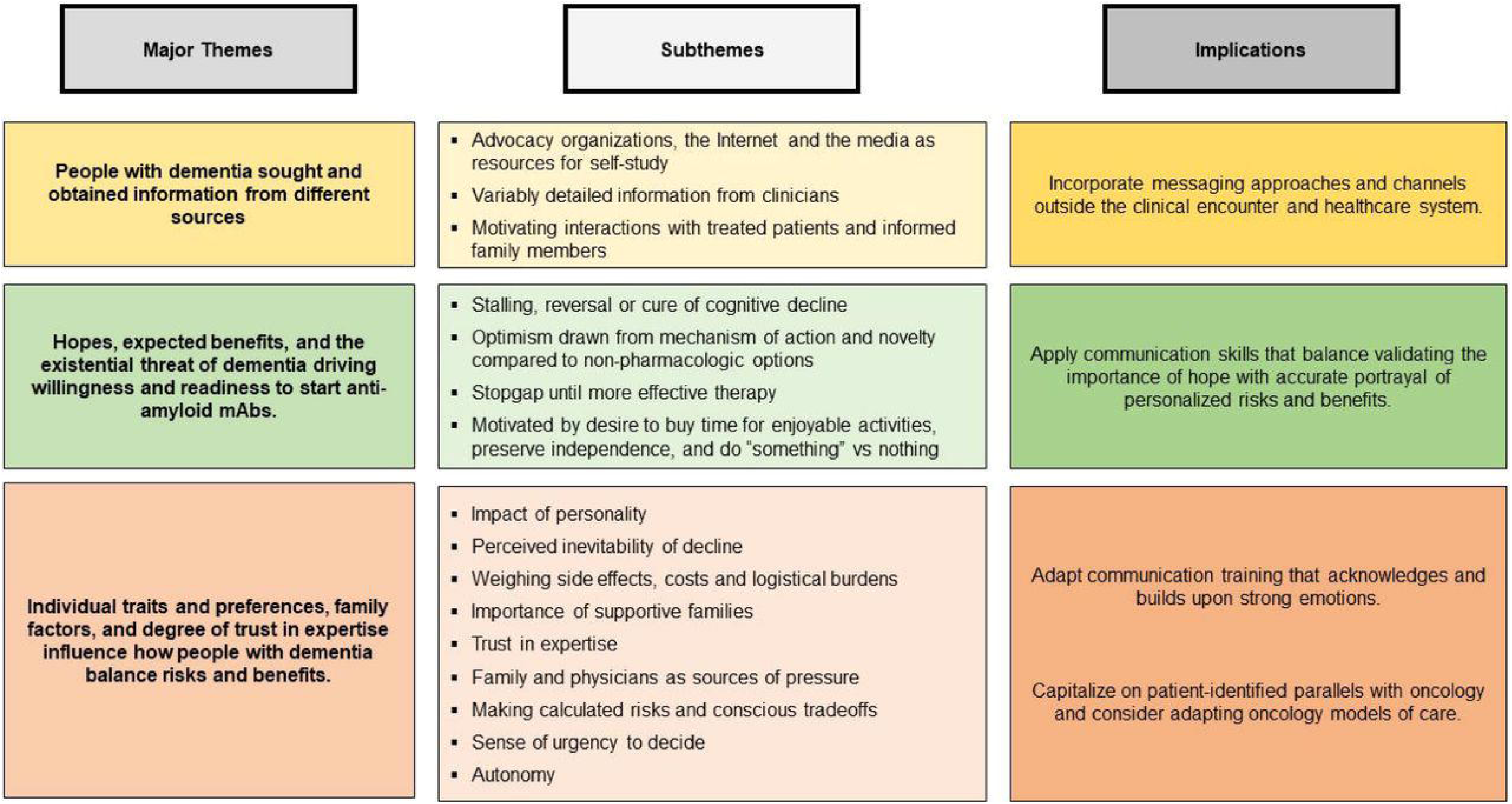
Major Themes, Subthemes and Implications Identified in People with AD Considering Anti-Amyloid Monoclonal Antibody Treatment.

**Table 2.**
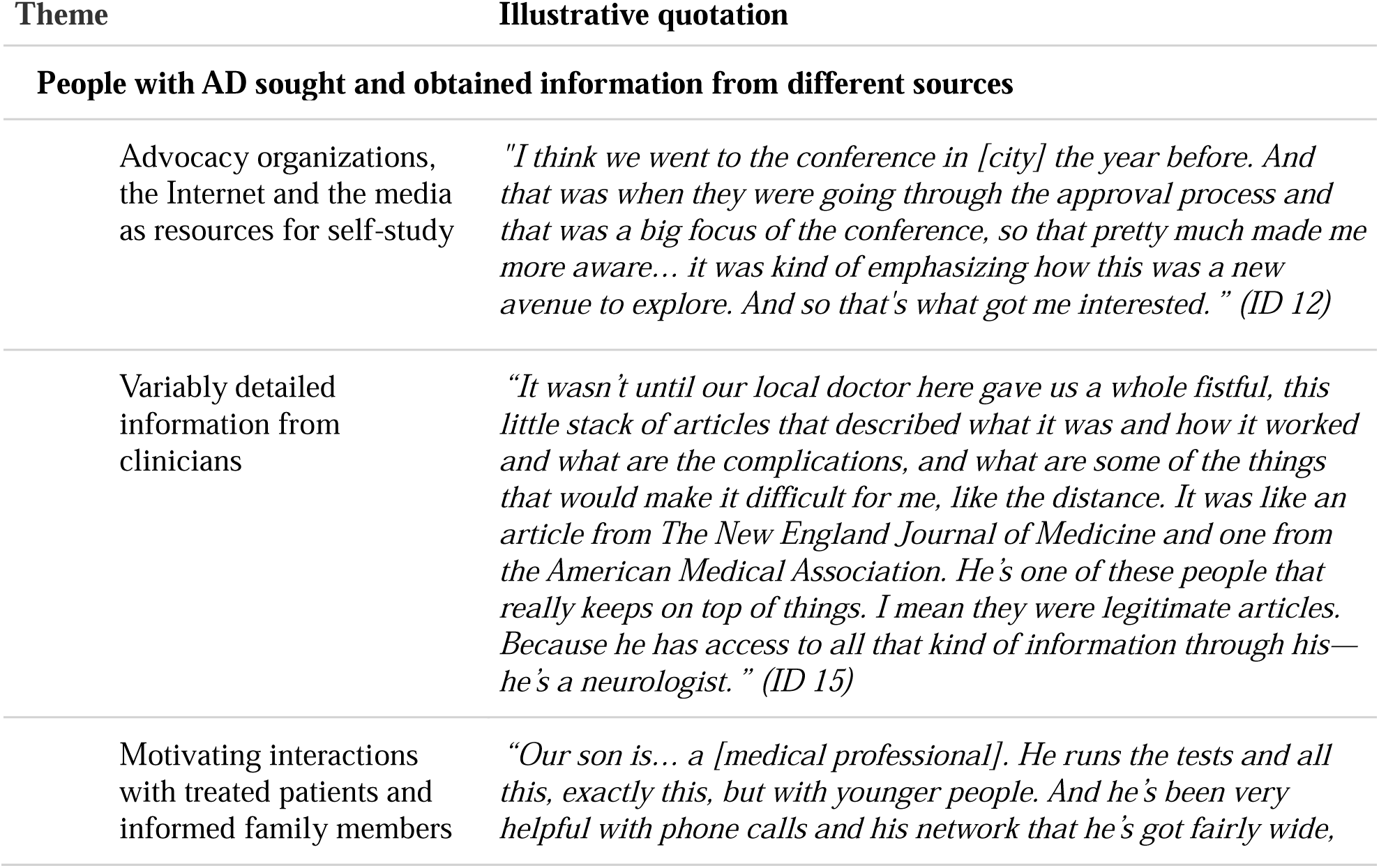
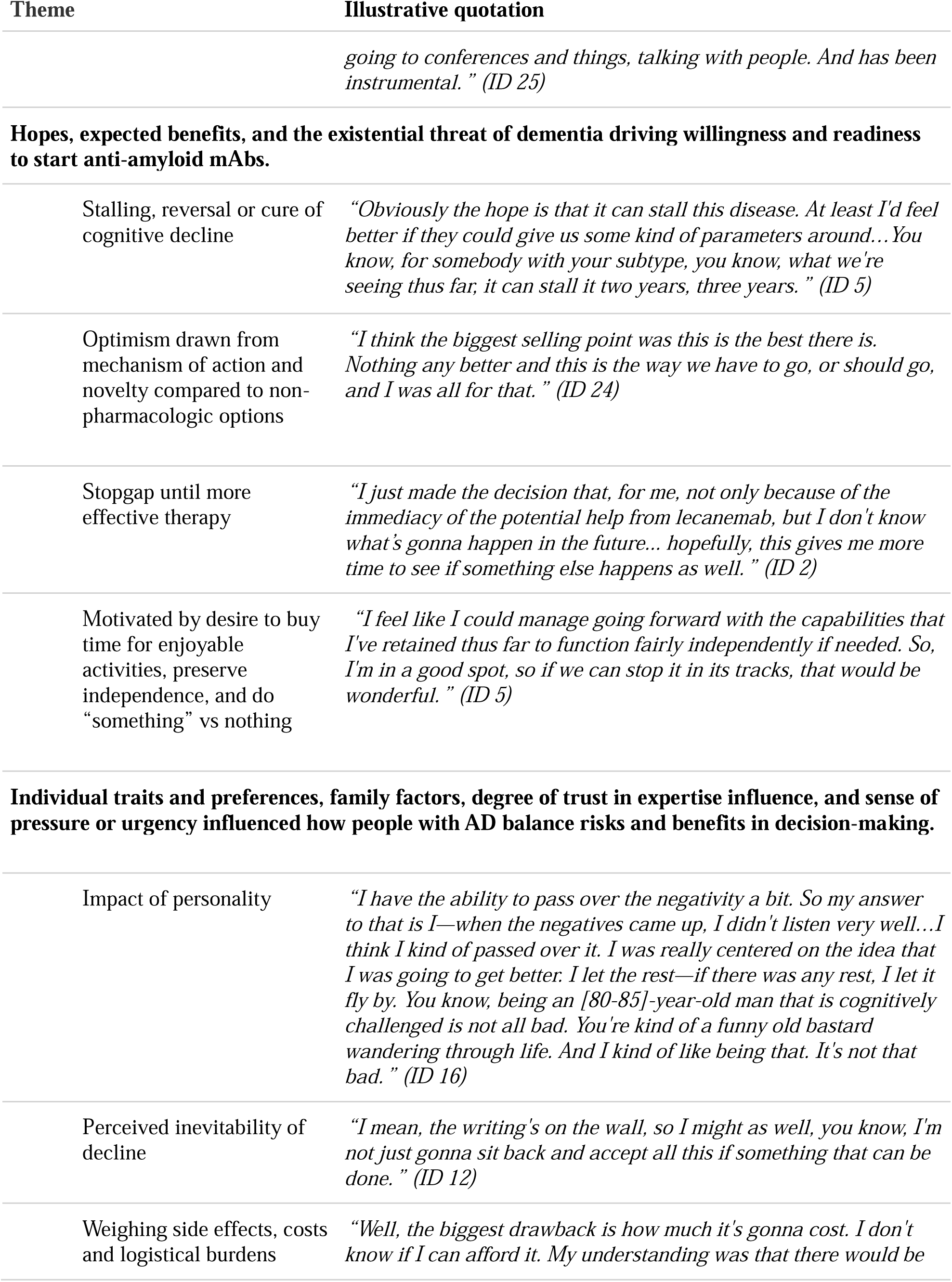
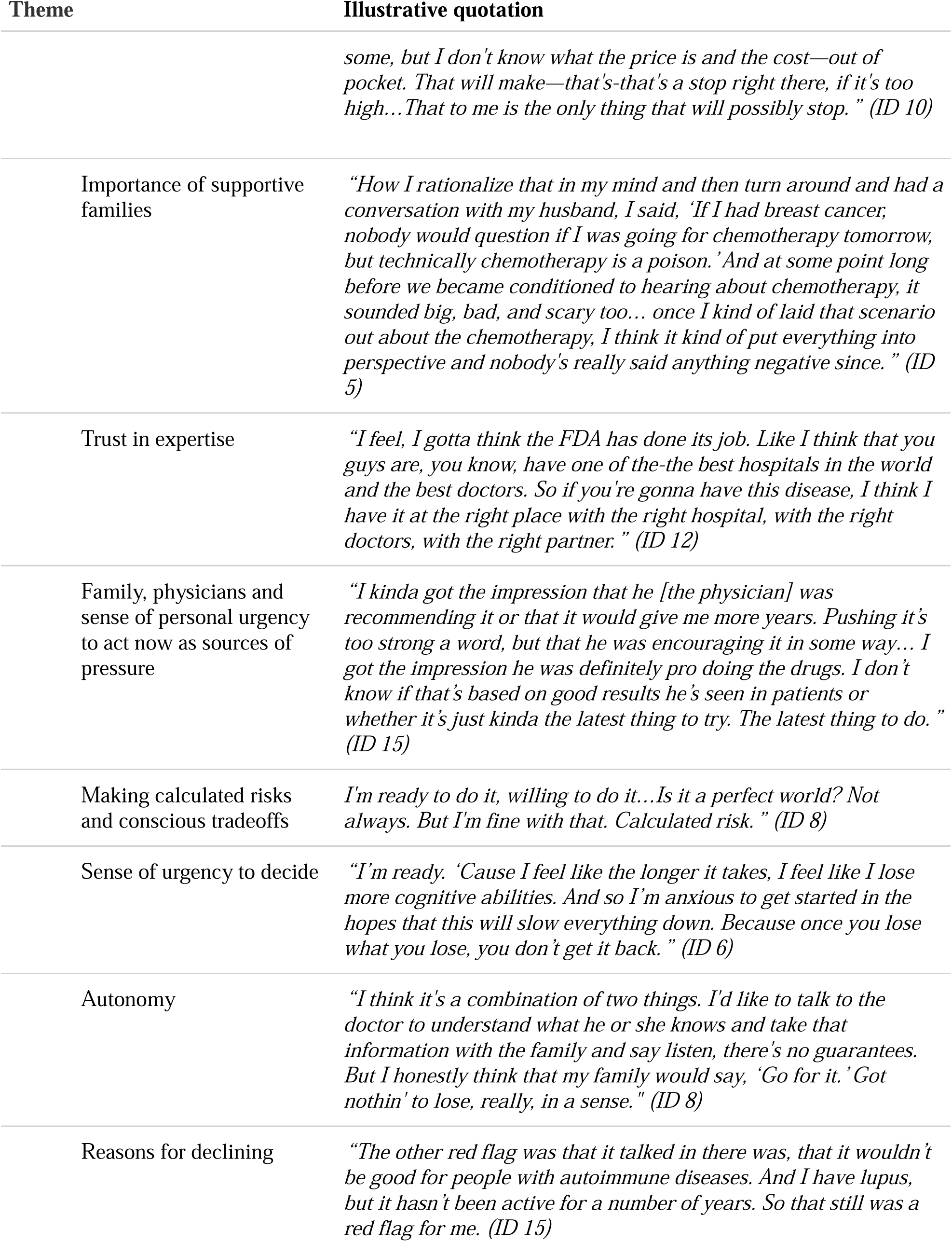
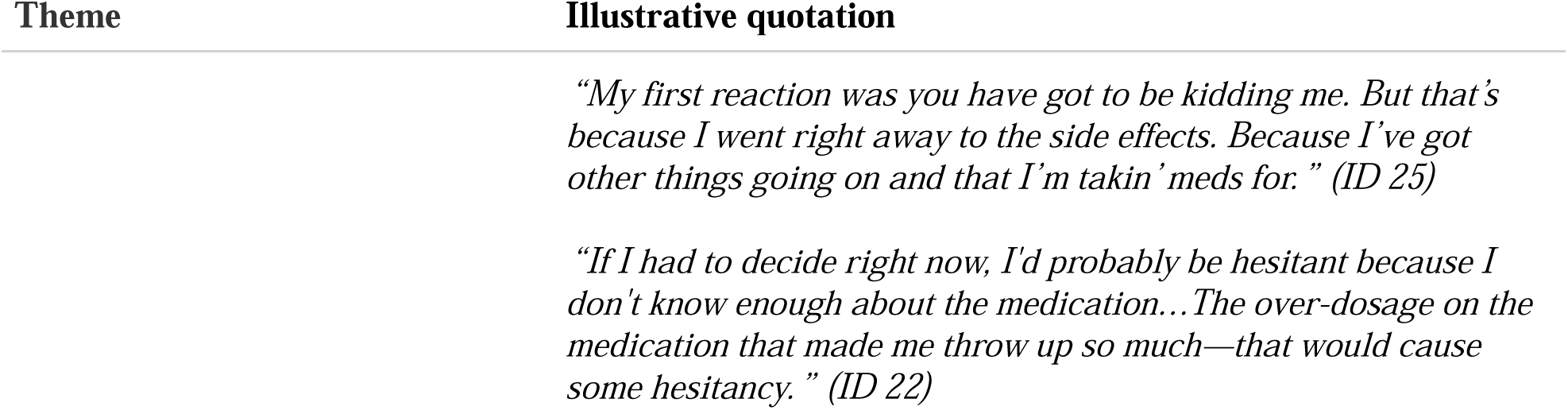
Additional Illustrative Quotations from People with AD by Theme.

### People with AD sought and obtained information from different sources

Nearly all respondents reported self-directed information gathering. In addition to the Internet and media, advocacy organizations played a prominent role in disseminating information and empowering patients.

> *“I’ve actually gone out and sought out information on my own…I was able to connect from the Alzheimer’s page to an article on it. And I basically printed that and then kind of read it in its entirety so that I could highlight areas of concern for myself.” (ID5)*

Most learned about anti-amyloid mAbs from their clinicians. Some learned from primary care providers—often the physician to whom they first presented with cognitive complaints—while others obtained information from subspecialty providers. Level of detail conveyed by clinicians varied, ranging from comprehensive to only a broad overview.

> *“You can do it over at our local hospital. I don’t really know much more than that…It just was, like, a couple of minutes or something. He just said it’d be an infusion, and so I asked him if that was—I don’t know. He didn’t really say much.” (ID 18)*

A few participants experienced motivating narratives from other people with AD who had received anti-amyloid mAbs. In other instances, information from family members with healthcare expertise played a role.

> *“…I met a gal who was a little younger than me who’s been getting the treatment and she really feels like it’s helped her quite a bit. She’s done very well on it, so that was another exciting thing.” (ID12)*

### Hopes, expected benefits, and the existential threat of dementia drive willingness and readiness to start lecanemab

Delaying progression of cognitive decline dominated the hopes for treatment. Some participants’ expectations exceeded the benefits seen in clinical trials. Several hoped for a cure, while others hoped that lecanemab would reverse memory loss or completely remove amyloid from the brain.

> *“[I hope] that they’ll come in in a year and say, ‘We can’t find any amyloid plaque—- in your brain. And that the tau tangles, or whatever they are, seem not to be there.’” (ID 9)*

Patient motivation to accept treatment came from the opportunities to a) gain more time feeling like themselves and doing enjoyable activities; b) have a plan to fight dementia rather than succumbing to the status quo; or c) preserve independence.

> *“I would like to be able to live a life and be able to speak and not get wound up in my mind…. This is the anxiety I’m hoping it can help not only just the hope, but it’s good to have a plan and, ‘This is what I’m gonna do.’ And ‘I’ve got good people around me.’ I feel good when that happens, I have to say.” (ID 8)*

Some people felt motivated knowing that they were receiving the “latest and greatest” compared to existing non-pharmacologic alternatives and that they were still cognitively able to assess the risks and benefits.

> *“I’m, you know, cognitively up with what people are saying to me and what I can read about or have my wife read to me about. So, I’m still pretty confident and —how do I put this? I’m worth saving.” (ID 22)*

Respondents also described lecanemab as a stopgap until the development of more effective therapies. This was coupled with anxiety about whether receiving lecanemab would preclude them from future treatments.

> *“The other thing that comes in is if you get on something, does it preclude the something better coming along? In the next two years you know, or whatever it is, that’s better. But because you’ve been on this or on this, you can’t do that. I don’t know how likely that is.” (ID 25)*

Some described feeling conflicted because they worried about tradeoffs (e.g., diminished quality of life from complications). Several raised concerns that lecanemab would have side effects like chemotherapy (e.g., nausea, hair loss) because both are infusions.

> *“It’s because, you know, I’ve had so much trouble with drug reactions in my life that I’ve tried as best I can to stay off drugs and stay away from drugs. So, I’m real—would be really concerned about, I’d rather have like two good years with as clear a mind as I can have, rather than having three years, but being so affected by the drugs that I’m taking that it wouldn’t be worth the extra year.” (ID 11)*

> *Individual traits and preferences, family factors, degree of trust in expertise influence, and sense of pressure or urgency influenced how people with AD balance risks and benefits in decision-making*.

When asked to consider risks and benefits, some noted that their approach was rooted in their personal tendency to “look past the negatives”, rather than the specifics of anti-amyloid mAbs. For others, the perceived inevitability of decline without intervention helped them decide to accept treatment at any cost.

> *“What do I have to lose? Is basically the way I look at it. I mean, even if it does cause problems for me, I’m headed that way already. I’m not—might be a different kind of problem. I don’t care.” (ID 10)*

The complication of ARIA-H weighed on most participants’ minds. Although people with AD wanted more information on their individual risk, they felt that bleeding risk was not high and surmountable. Very few mentioned death as a potential outcome.

> *“I just know that there’s things that could go bad, and I’m ready for that. I’d rather be able to get onto the drug and… I’d rather be able⍰ on it. At least try it, and… if I die, I die.” (ID 17)*

Costs, insurance coverage, and logistical hurdles also influenced willingness to pursue treatment. Some felt that infusions every two weeks would not affect their lifestyle, while others found this disruptive. People with AD likened the logistics of anti-amyloid mAbs to receiving chemotherapy for cancer.

> *“It also goes with a medication that caused you to lose what you already have as far as your physicalness, your aliveness, those kinds of things. So if it’s a medication that’s going to maybe give you longer memory, but less life, that would make a difference.” (ID 25)*

Family also influenced how people with AD weighed side effects and costs. Some described family members as being very supportive, while others described hesitation. Those whose desire for treatment conflicted with family members reminded them of their autonomy despite their declining cognition and again drew comparisons to cancer to address concerns. A few equated forgoing treatment with unnecessarily burdening family.

> *“My husband has been wonderful about it. I think he has more worries about the risks than I do, partly because he’s like, ‘I’d rather have you even if you don’t know who I am anymore. I’d rather have you as long as I can.’” (ID2)*

People with AD voiced trust in expertise and felt that the medical team took their individualized risk and benefit into account. Faith in the drug approval process and their healthcare system also influenced decision-making.

> *“I know that no doctor is perfect. I do honestly believe that they know a lot more about this than I do—in many ways. And I trust that they’re very, very good. And that’s the only way I know how to answer…I don’t even think about it because I trust it. Unless something went wrong, then it might change… It’s like I say, I feel like something happens that makes me lose that trust. That’s hard to get back.” (ID 10)*

People with AD described a sense of pressure to accept treatment. Pressure came from family members, clinicians, or a sense of personal urgency to act now (versus waiting until they may not have a choice).

> *“I suppose I could wait later, but I also feel like the damage that is happening, I know at some point—I don’t wanna get to the point where I don’t have the choice anymore, right?” (ID 2)*

When making a final treatment decision, a small proportion indicated that they would decide alone, and a small proportion would make a group decision with family. The majority described gathering input from physicians and family but ultimately felt that the decision was theirs to make.

> *“I will decide on my own—with agreement from the doctor. I’m certainly willing to listen to any concerns from my family member, but ultimately it is my body, my decision.” (ID 5)*

### Ideal future state of knowledge and decision-making

We also synthesized information that people with AD wanted to improve their knowledge and decision-making (**Table 3**).

**Table 3:**
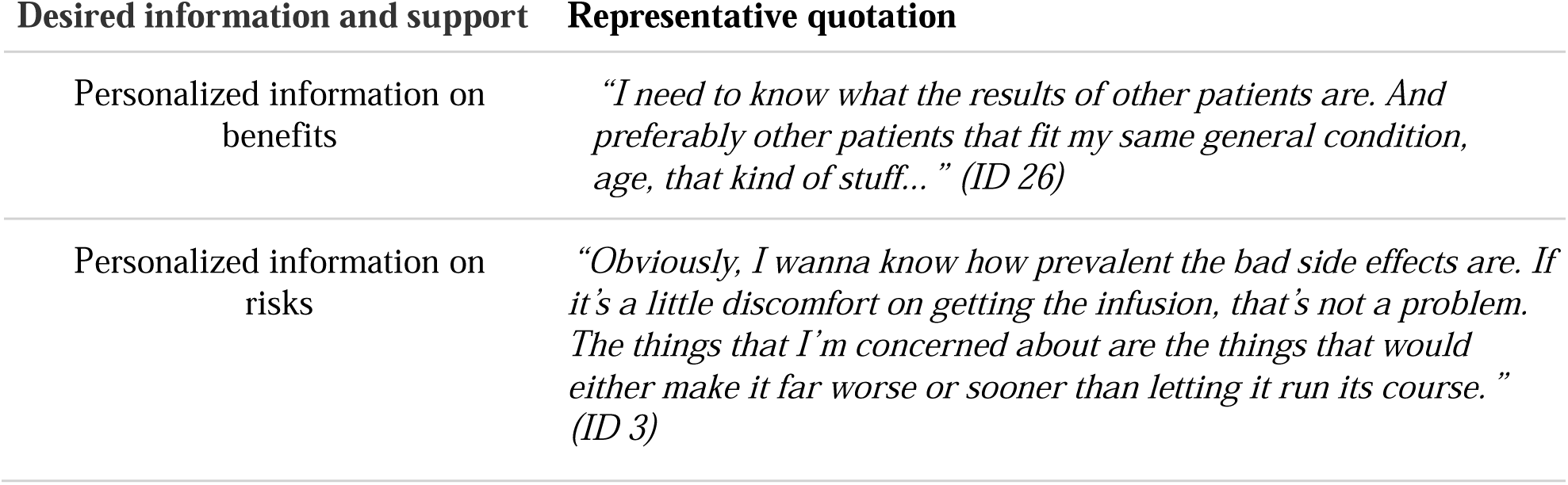
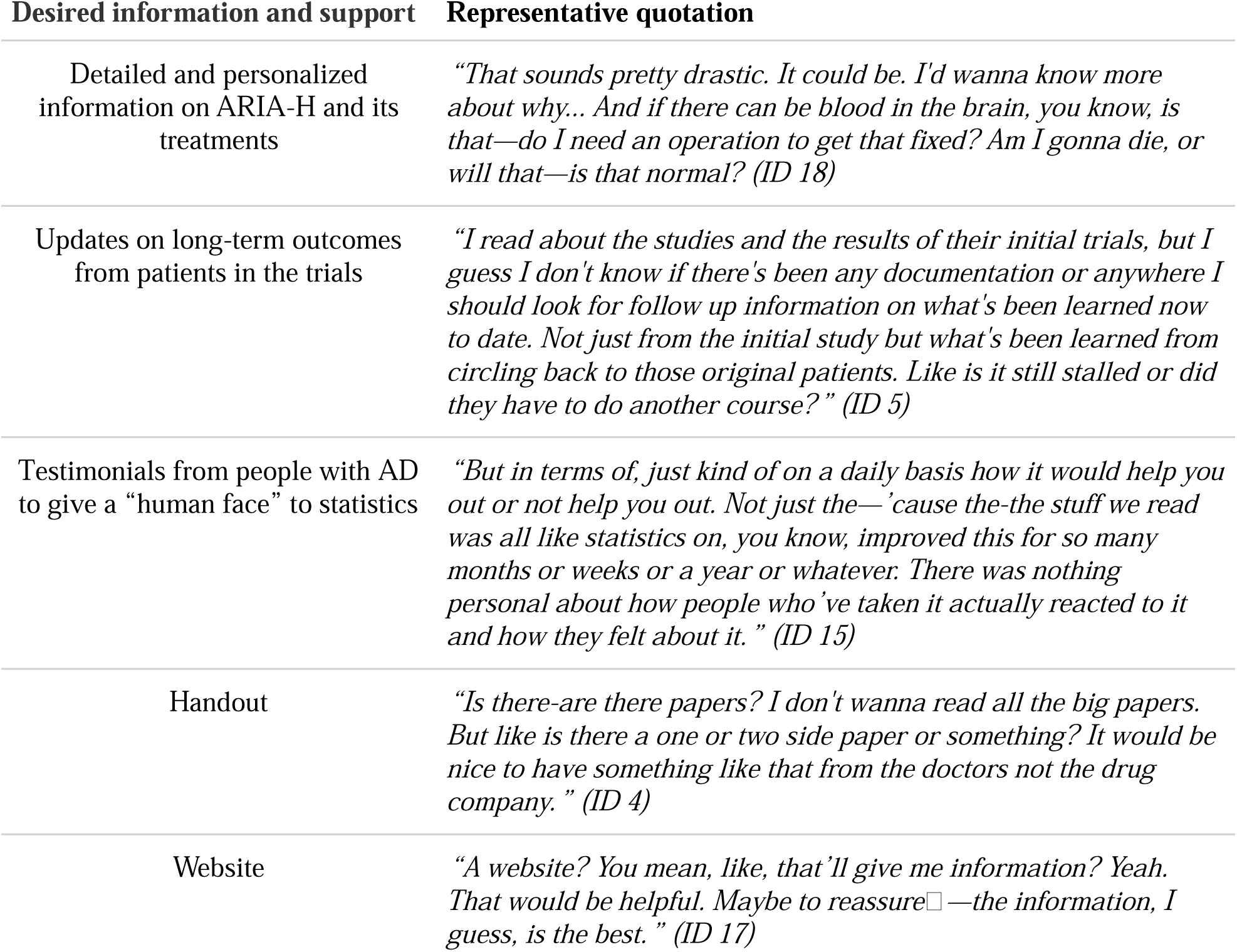
Desired information and support for anti-amyloid mAb treatment decisions.

| Desired information and support | Representative quotation |
| --- | --- |
| Personalized information on benefits | *“I need to know what the results of other patients are. And preferably other patients that fit my same general condition, age, that kind of stuff...” (ID 26)* |
| Personalized information on risks | *“Obviously, I wanna know how prevalent the bad side effects are. If it’s a little discomfort on getting the infusion, that’s not a problem. The things that I’m concerned about are the things that would either make it far worse or sooner than letting it run its course.” (ID 3)* |
| Detailed and personalized information on ARIA-H and its treatments | *“That sounds pretty drastic. It could be. I'd wanna know more about why... And if there can be blood in the brain, you know, is that—do I need an operation to get that fixed? Am I gonna die, or will that—is that normal?” (ID 18)* |
| Updates on long-term outcomes from patients in the trials | *“I read about the studies and the results of their initial trials, but I guess I don't know if there's been any documentation or anywhere I should look for follow up information on what's been learned now to date. Not just from the initial study but what's been learned from circling back to those original patients. Like is it still stalled or did they have to do another course?” (ID 5)* |
| Testimonials from people with AD to give a “human face” to statistics | *“But in terms of, just kind of on a daily basis how it would help you out or not help you out. Not just the—'cause the-the stuff we read was all like statistics on, you know, improved this for so many months or weeks or a year or whatever. There was nothing personal about how people who've taken it actually reacted to it and how they felt about it.” (ID 15)* |
| Handout | *“Is there-are there papers? I don't wanna read all the big papers. But like is there a one or two side paper or something? It would be nice to have something like that from the doctors not the drug company.” (ID 4)* |
| Website | *“A website? You mean, like, that'll give me information? Yeah. That would be helpful. Maybe to reassure□—the information, I guess, is the best.” (ID 17)* |

## Discussion

To our knowledge, this is the first study evaluating how people with AD understand and deliberate about anti-amyloid mAb treatment (**Figure 2**). People with AD sought out information to supplement knowledge from their physicians. They hoped that state-of-the-art treatment would forestall inexorable cognitive decline and bridge to more effective treatments. They were motivated by taking action that could buy more time, rather than passively letting nature take its course. Diminished quality of life due to side effects, costs and burdens of treatment were concerns. How people with AD balanced these tradeoffs was determined by personality, family, and confidence in expertise.

Our findings help clarify a new stage in the dementia disease trajectory: decisions about disease-modifying therapy. Before effective therapies were developed, qualitative studies in people with AD focused on experiences and end-of-life care. Our findings have some similarities, including a shared desire for greater guidance from clinicians on expected disease course. Distinct from our findings, participants in prior studies focused on behavioral (versus pharmacologic) interventions and the need for more palliative care.^18–21^ Research examining decision-making for people with dementia focuses on moving to a nursing home and advance care planning. Some key facets overlap with our findings, like variation in perceptions of risks and benefits, emotional complexity, and desire for autonomy. ^22,23^ In a recent systematic review of 13 qualitative studies, true decisional autonomy was rare and confined to minor decisions, whereas it was nearly universal in our population with mild symptoms.^24^

Our study participants were predominantly White, although they exhibited more educational, income and geographic diversity. During the recruitment period, 97% of patients seen at the two study sites’ memory care clinics were White, which is in line with the population characteristics at memory care clinics nationally.^25^ It is also consistent with a recent analysis of real-world rates of lecanemab prescription between January 2023 and February 2024, which revealed that 80% of patients receiving lecanemab were White.^26^ Decades of social science research have empirically demonstrated that medical breakthroughs, even when beneficial to health, exacerbate health inequalities.^27–29^ Treatments that are expensive, complicated or time-consuming—like anti-amyloid mAbs—are more likely to create disparities. This is because those with socioeconomic advantages (educational, financial, social) will employ them to overcome these barriers and obtain access to new treatments. The lack of diversity in patients seen in these two memory care clinics and among participants in our study, therefore, suggests a recapitulation of this phenomenon and an imperative to intervene. Subsequent studies will examine whether our findings apply in settings with greater diversity that reflect all people with AD. As anti-amyloid mAb science continues to advance, concerted efforts to reduce cost and complexity of care should promote broad access and reduce disparities for people with AD.^29^

These results also reveal the sensitivity and skill with which clinicians caring for people with dementia must approach conversations about treatment. Our results inform how communication strategies could be adapted for this need. Some participants had misconceptions about lecanemab’s benefits, and potential side effects were considered minor. Given the gravity of a dementia diagnosis, expressions of “unrealistic hope” may reflect normal coping or a lack of cognitive understanding.^30,31^ Some clinicians may be prone to overstating benefits and minimizing harms, while others focus on potential adverse sequelae.^32^ Our results also underscore the salience and complexity of emotions in treatment decisions for people with AD, such as grief over loss of identity and worries about burdening family. Evidence-based communication training could address these needs, including sharing personalized risks and benefits, naming uncertainty, using pictorials, creating space for questions, partnering in decision-making, and using empathy.^33–37^

Our findings highlight that promoting informed decisions about anti-amyloid mAbs would benefit from incorporating messaging channels outside the clinic. External resources were important for people with AD. Heightened media attention to anti-amyloid mAbs may have led to passive exposure.^38–40^ Alternative resources may also have filled a vacuum while awaiting subspecialty appointments or when incomplete information was provided by clinicians, a phenomenon seen previously in dementia.^18^ Interestingly, no people with AD reported hearing about lecanemab through direct-to-consumer marketing, which is both common and can distort decision-making.^41,42^ Given patients’ multiple streams of information, interventions to provide education and decision-making support should use various modalities and start before and continue after interaction with the healthcare system.

Patient-drawn parallels between lecanemab and chemotherapy raise potential opportunities to learn from both the successes and failures of oncology care models. This comparison is apt given the rapid transformation of AD care into a landscape not unlike oncology care. People with dementia who might previously have felt stigmatized and hopeless about their diagnosis may now identify with cancer patients who often have access to effective therapies and are valorized for their bravery.^43^ Clinicians caring for people with memory loss must now add responsibilities that are common among oncologists like in-depth side effect monitoring.^44^ Healthcare systems must now provide services common in oncology care, including helping people with dementia navigate complicated visit schedules. Lessons learned about the limitations of oncology care models can be applied, such as the potential for financial considerations to affect treatment decisions and the importance of eliciting patients’ values and aligning care to meet them.^45,46^ Successful aspects ofoncology models could be adapted and their pitfalls avoided to support people with dementia, clinicians and healthcare systems in making informed, cost-conscious and goal-concordant decisions about anti-amyloid mAbs.^47,48^

### Limitations

Although we reached data saturation, our study may not capture the full spectrum of patient perspectives. Aspects of decision-making overlapped with those in previous studies in dementia, suggesting broader relevance.^18–21^Our participants had some of the earliest access to anti-amyloid mAbs. These early adopters may not reflect all people with AD. For example, we identified consistent themes of self-efficacy, autonomy, and eagerness to start. Our findings could simply be interpreted as natural enthusiasm in response to relief from a devastating—and previously unmodifiable—diagnosis. Additionally, participants were recruited from memory care clinics at academic medical centers who may differ from those in community settings or with limited access to specialized care. As noted above, our sample lacked racial and ethnic diversity so may not be generalizable, and future studies targeting underrepresented populations will advance our understanding of shared decision-making. Future studies should also examine how patients’ cognitive status affects their perception of risks and benefits and decision-making. Patients’ views on treatment can evolve with time; longitudinal studies examining changes in decision-making would also offer important insights.

### Conclusions

In this first qualitative study of front-line people with dementia considering treatment with anti-amyloid mAbs, we found variation in hopes for treatment, information sought and obtained, and contextual factors in decision-making, supporting the need for an individualized approach. Identifying these features can help identify targets for interventions to improve individualized decision-making for anti-amyloid mAb treatment.

## Supporting information

Appendix 1 COREQ Checklist

Appendix 2 Master Document Thematic Statements

## Acknowledgements

We appreciate the time and expertise of our study participants who contributed data.

## Author Contributions

*Anna L Parks, MD* (Conceptualization; Data curation; Formal analysis; Funding acquisition; Investigation; Methodology; Project administration; Resources; Supervision; Validation; Writing – original draft)

*Ayush Thacker, BS* (Data curation; Formal analysis; Methodology; Project administration; Writing – review & editing)

*Daniel Dohan, PHD* (Formal analysis; Investigation; Methodology; Resources; Supervision; Writing – review & editing)

*Liliana A Ramirez Gomez, MD* (Conceptualization; Investigation; Writing – review & editing)

*Christine S Ritchie, MD, MSPH* (Resources; Supervision; Writing – review & editing)

*Joanna Paladino, MD* (Conceptualization; Data curation; Formal analysis; Investigation; Methodology; Writing – original draft)

*Sachin J Shah, MD, MPH* (Conceptualization; Formal analysis; Funding acquisition; Methodology; Writing – original draft)

## Statements and Declarations

### Ethical considerations

The institutional review boards of participating institutions approved this study as exempt.

### Consent to participate

Study participants completed verbal informed consent for participation.

### Consent for publication

Not applicable

### Declaration of conflicting interests

The author(s) declared no potential conflicts of interest with respect to the research, authorship, and/or publication of this article.

### Funding statement

Dr. Shah reports research funding from NIA (K76AG074919). Dr. Parks reports research funding from NIA (K76AG083304).

### Data availability

The data supporting the findings of this study are available within the article and/or its supplemental material

## Supplement

Appendix 1 COREQ checklist

Appendix 2 Master coded thematic statements

## Notes

### Competing Interest Statement

The authors have declared no competing interest.

### Funding Statement

This study was funded by the National Institute on Aging (K76AG074919).

### Author Declarations

The Institutional Review Board of Mass General Brigham gave ethical approval for this work

### Summary of Updates

We have updated the methods of recruitment and enrollment to clarify our sample size

