## Appendix 2 Master Document Thematic Statements for "A Qualitative Study of People with Alzheimer’s Disease in a Memory Clinic Considering Lecanemab Treatment"

### *Patients sought and obtained information* from different sources, including advocacy organizations, the Internet, and clinicians.

Sought out Alzheimer’s related media/advocacy

- I've actually gone out and sought out information on my own. So, um, from—I was able to connect from the Alzheimer's page to an article on it. Um, and I basically printed that and then kind of read it in its entirety, um, so that I could highlight areas of concern for myself (ID5)
- “my [relative] and I are both pretty veracious in-in reading and trying to get all the information that we can. Um, I will say, uh, we’ve done a lot of it ourselves. It’s, uh—and some of that is just because that’s who we are. Um, I would say, uh—and I’m—this isn’t a criticism, per s-per se, because-because we were doing so much, maybe I didn’t, uh, really ask all that much. But I would say we haven’t actually gotten a ton of information from the hospital.” (ID 2)
- Well, <Name> really does most of the, uh, processing and information, but I just, you know, what I see on the internet and on the AL'S web-website, uh, you know, I-I was probably even aware that it was in the pipeline, maybe, uh, as long as two years ago, right? A year and a half, two years ago. (ID 12)
- *“*Learned about it in-in [city] and everything. So, uh—and so, that was *[distorted audio 04:11]* and the thing before that, um, I was thinking that it’s like—all that time e-when they were talking about the drug-the new drugs there—they’re saying-everybody was saying they were-we're running out of time. You know? So-so—'cause I have-I-I have Alzheimer’s, and, um, I had-I guess I had three years now and—almost three. And, um, you know, so everyone was fighting. (ID 4)
- Well, I think we went to the, uh, conference in [city] not this past year, but the year before. And that was when they were going through the, uh, approval process and that was a big focus of the conference, so that pretty much made me—- more aware, yeah, there was a presentation and, uh—- it was kind of emphasizing how this was a-a new-new, uh, avenue to explore. And so that's what got me interested.(ID 12)
- Um, probably in-in what I read, obviously it talks about like the, um, concerns with ARIA and I do know that I am, uh, not an AD, what is it? Oh, we-I'm not a APOE e4 carrier. Um, so that made me feel a little bit better about, you know, potentially less risk for side effects. (ID 5)
- “I, uh, am aware of the side effects.” They tell me that, uh, I have, um, I do not have the genetic indicators that indicate higher risk. (ID 9).

Biotech/Investments space

- I work, um, in investments, and so in the biotech, um, stuff we're always looking, um, at, uh, what's coming. And, um, and then with the—we're—we're just—actually, probably the Internet. And, um, I saw it. And then the FDA approved the Alzheimer's drug in July, I think. but then when we heard about—the Leqembi We thought, "Oh." "We should know about this." "And everybody should know about it." And I know there are other areas in, uh, the US that—that have it. And—and are using it. And—and, uh, I guess it's—I don't know if it's, uh, if it's blessed, you know, that—that it's perfect. (ID 8)
- Um, my-my source of information, if-if you're in the <City> area, it's-it's heavily investment-oriented and it's*—*that's a lot of what, uh, drives the-the news. I don't know if you can see this or not, but news for the product is there. And, uh, so anyway, that's, uh, an important source of information is the investment community. And <Organization> is-is a big player out here. (ID 11)

General Media

- Oh, and 60 minutes. I did see the piece in 60 minutes. (ID 5)
- I saw the drug in—I think it was AARP. I thought—I think that was it… So, um, I think it’s a drug that could slow the progression of the brain—um, of the brain, I guess, or something like that. (ID 17)
- “A few weeks ago I read in either the New York Times or someplace that drug” (ID 9)
- Uh-uh, the most helpful information was the ARRP-AARP article. That was probably the best one, but, uh-uh, we got to talk with, uh, <doctor> for a little bit about it, and she filled us in pretty good, so. (ID 22)
- Oh goodness. I just went online and-and, uh, and I think probably what I typed in was, uh, the word.Yeah. And it—- and it—and then I read what it had to say. And of course it had, you know, side effects—- as part of that. (ID 25)

Providers

- As far as what I've heard from providers, just that, um, it is out there. Um, it's showing very promising results. Um, and when I saw, uh, <Name> at <Hospital>, he felt that I was a good candidate for it. I did go out and look at it through my health insurance, um, like a benefit site just to see. I wanted to see if it was something that was covered (ID 5)
- and, um, so, the first time—like once I got my diagnosis, like, you know, I got the-the, uh, the drugs that usually come. But that-but it’s like-it might help...it a little bit—best way and save me as fast as you can and all that stuff. Um, and so, she she had a list for me, and I had-I think there was 14 things on the list, and the last one was to...[see] if I'm a-I'm-I-if I would be a bleeder....I know my [relative] had emailed her (ID 4)
- Only from Dr—I can't remember her name right now. But—I can't remember it.....I don't remember all the details about it other than I was like I would like to give this a try. (ID 10)
- my [medical professional] was very aggressive in looking into things and-and particularly was the one who pushed about doing the testing for the APO, um, 4. So, we knew that and that that was much more of a-an indictive of that. Maybe that’s what was going on with me. So, I had a good doctor, but he was the first to say, “I’m a generalist. You know, I’m a”—*[laughter]* and there just aren’t any services.” (ID 2)
- Well, um, our [medical professional] here in <City>, who’s also a real close friend, gave us a number of articles on, um, the test results and the, um, you know, the drawbacks and the-and the problems. And, you know, what needs to, uh, happen in connection with that. So, um, yeah, I read through’ em last night and I think<Name>readthrough ’em the other day. So we know a little bit. Um, you know it's a little—still a little confusing for me. But I think I understood a lot of it. (ID 15)
- “it wasn’t until, um, our local doctor here gave us a whole fistful, this little stack of articles that described what-what it was and how it worked and, you know, what are the complications, some of the compli—possible complications and what-what are some of the things that, you know, would, um, make it difficult for me, like the distance, you know. Yeah. It was like an article from The New England Journal of Medicine and one from the American Medical Association. **He’s one of these people that really keeps on top of things.** Um, I mean they were legitimate articles. ’Cause he’s, um, you know, he has access to all that kind of information through his—he’s a [medical professional] (ID 15)
- Well, as I understand it, it's an injection that I take every two weeks. Well, we met over a couple of, uh, pictures of my brain, and it was explained to me what this will do, which should—it was my understanding—which **should bring me back closer to where I was, or at least not losing that ability;** and that there was going to be six weeks to a year—**hadn't quite decided yet how long I was going to go with it.** But the way it was explained to me is—is that it would take years and years and years to get back to where I am, which is the good part. Probably wouldn't outlive it. Like the doctor was telling me, he said, "Hell, I don't think I'll outlive it."…yeah, so that was very encouraging. (ID 16)
- You can do it over at our local hospital. I-I don't really know much more than that.It was with—when I was with Dr. <Name> at the <Institution>. Um, I don't really recall. It just was, like, a couple of minutes or somethin'. He just said it'd be an infusion, and, uh, so I asked him if that was—I don't know.He didn't really say much. (ID 18)
- He told me that it was a couple of, um—couple of times a month, and I didn't need a, um—like, I guess it's just inner vein—intravenous 03:06 that-that's supposed to help my brain or something. (ID 18)
- And technically, he’s actually—you know, he has actually given—the first doctor has given me a little bit of an overview of what it is and why-why it’s something we wanna—he wants us to consider, or he think we should consider…But-but knowing the actual details about it, no. (ID 3)
- Uh, sure. Um, he basically—as I understand it, there is a study has come out that shows that this drug, when taken, uh, does reduce the amyloid buildup in about half the cases. And that-that amyloid buildup is what they think contributes significantly to the memory loss and other problems with Alzheimer’s. Um, he did-he did mention that there are some side effects, um, have occurred, some very bad side effects have occurred. Um, but that’s really about it. (ID 3)
- Um, I think he just gave me the-the-the short, uh, brain bleed as the worst one. Which I think kinda speaks for itself. (ID 3)
- It was a Zoom call. No, I think Zoom calls are wonderful—- for almost everything. (ID 3)
- Um, really, the only one I’ve talked to is my doctor. Not really, no, not that I remember anyway. Um, just that, you know, I’d have to come every other week for the treatment for quite some time. I don’t remember the amount of time, but quite a bit time. (ID 6)
- We've kind of looked into it a little bit, but, um, uh, actually, a doctor here in, uh, <hospital> contacted us- and said that we might be good candidates. (ID 22)
- We’ve met with many, many people over the last six months on this.Uh, numerous results and tests. I mean sessions with doctors. And explanations of—with several different types of doctors. (ID 25)
- Um, well once, uh, we realized there-there was something going on, uh, we checked with prime—my [medical professional], and, uh, she suggested we go to the <Organization>.
- Um, and that's what we did, and, uh, that's where we have been getting all the information that, uh, led us to Dr. <Name> and, uh, his group. (ID 26)

Patient who had a significant impact

- I guess I did hear about it from one other person. I participate in a support group on Mondays of people with Alzheimer's. And there was a gentleman in that group. Um, he took it for a year and he had positive-positive results. He was actually, um, an attorney who was, um, just going back to work on a consulting basis after having not been working because of his disease progression. (ID 5)
- We met—we went up to, uh, <City> to do a—I don't know, whenever they had lobbying. They seem to send us so, um, I-I **met a gal who was a little younger than me who's been getting the treatment and she really feels like it's helped her quite a bit. She's done very well on it, so that was another exciting thing..... I**-I get these fillings of deja vu, so I go up to her and I go, do you ever get a feeling? Oh yeah, I get that. And, you know, her [family member] looking at her like, she's nuts. And my [loved one] looking at me like, I'm nuts. And oh, she just gave me the bird too. So-so it was-it was kind of nice to see that somebody else has-has experienced that. (ID 12)
- [I have a friend] who had, um, um, relatives who, um, were not eligible. They were too old when lecanemab first came out. Uh, but I’ve talked to—with folks who have gone through the processes of having the disease. Um, and, you know, and a lot of conversations *[laughter]* with my family, right, with my kids and everything about there are risks. And because I have the double APO gene, I have a little more risk. Um, but at the moment, we just decided I’m so young that, um, quite frankly, I-I don’t—I’m not ready to give up yet. *[Laughter]* So, we just, uh, made the decision that we’ll take it one step at a time if any risk factors come up in terms of the potential bleeding, *et cetera*. (ID 2)

Gender differences

- Saw on the statistics that-that we saw in those papers where, um, the—how there was a difference between me and women in terms of effectiveness. And it was a lot less for women. Um, a lot less, um, positive outcomes for women. (ID 15)

Awareness of other treatment options

- , certainly looking at some of the, um, you know, exercises that-that you can do on paper and, you know, kinda do those things. But, um, what we kinda determined was, particularly because I’m so young, *[laughter]* that, um, you know, this isn’t a matter of, can I do, you know, logic problems and things like that and just try to stay a little bit smarter for a while? Um, the question was—is, how can we maximize clarity, you know, so that I can continue to write books or whatever else I wanna do and-and enjoy that? (ID 2)
- I don’t believe there are any other treatment opt right—options—- at this point. I mean-I mean, they’ve—the doctors have come up with things I should be doing like exercise and sleep and eating well and all that, but that kind of—Yeah. So, we build Legos and do puzzles, which are interesting, (ID 3)

More info needed:

Differences in subtypes of ADRD

- I would've wished that it may be included more information with regards to, um, like the subtypes of Alzheimer's and what, um, maybe what the studies have shown in regards to that, if it helps some groups more than others. (ID 5)

Handout

- I-I would've probably appreciated like a handout on it versus kind of having to track down the information myself. (ID 5)
- Um, I guess I should find out the other downsides for it.Is there-are there papers? I-well, I don't wanna do-I don't wanna read all the big pi-pi—all the papers. But like is there-is there a, um, one or two paper side or something-anything? it would be nice to have something like that (website) from the doctors (not the drug company) (ID 4)
- I’d like to have the handout, and then, uh, somebody available to answer the questions that we would have.And-and I would like to-to locate the source. (ID 20)
- Yeah, I mean, I*—*if you write it and send it, I'll have it. And *[laughter]*, I do read 'em, uh, I am, you know, a little bit light in the knowledge department, so I-I need to, you know, get-get more deeply involved in it and whether you have a*—*some other*—*another avenue to get-to get to the end result. I-I*—*I'd-I'd love to see it, could be written. It could be, you know, media. (ID 11)

Website

- Well, maybe. Uh, that could be. A website? You mean, like, that’ll give me information? Yeah. That would be. Yeah. That’d be helpful. Maybe to r-reinsure—The information, I guess, is the best. (ID 17)

Showcasing updates from patients in the trial

- “Maybe if-if—I read about the studies and the results of their initial trials but I guess I don't know if there's been any documentation or anywhere I should look for follow up information on, um, what's-what's been learned now to date. Not just from the initial study but what's been learned, um, from circling back to those original patients. Like is it still stalled or did they have to re—you know, do another course (ID 5)
- I haven’t had an opportunity to speak with anybody who’s actually take-taken the pro-protocols. Um, so, that—I don’t—that’s the only piece kind of missing in terms of-of what’s going on there. (ID2)
- maybe more just the simple information of g—is there a way to have testimonials from people who have now gone through the first tests? Or, you know, something like that to really understand more of the human effect of what people went through taking the-the medicine. I really don’t have a lot of information on that yet. (ID2)
- “I don't know if there’s, you know, people who have gone through the protocols, if they ever do any kind of presentations themselves. That would be great to hear and listen. Um, I-I know the—it’s also new. I realize that it’s not, like, you know, cancer survivors have all kinds of, you know, groups outside or something like that. Um, it’s something that if, um—you know, if that’s something that is available or as it starts to become available or if it's something that” (ID 2)
- I guess, you know,I-I would be real interested in hearing from, uh ,people who’ve actually taken the drug to see how it affected their daily lives,um, after-after an infusion or before an infusion. Um, and if they,youknow, felt better or they—their minds were clearer or—you know, I don’t know. I guess that’s thekind of information, you know, just actually from, you know, patients who’ve taken it. And what-what are some of the potential drawbacks to that.Or if it makes ’em feel great for a couple of weeks. I don’t know (ID 15)
- **Um, but in terms of, um, which is great, you know. But in terms of, um, just kind of on a daily basis how it would help you out or, um, not help you out. Not just the—’cause the-the stuff we read was all like statistics on, you know, improved this for so many months or weeks or a year or whatever. Um, but it’s there’s no—nothing—there was nothing personal about how people who’ve taken it actually reacted to it and how they felt about it. (ID 15)**
- Maybe I mentioned this already here, my memory is not great. Um, but, you know, there was no place in those articles that actually talked about how the-how the patients feeled, felt, after, you know, after the treatments. And-and, uh, you know, over a period of time. So I really didn’t get a good idea of, you know, people saying yeah, do this, it’s wonderful. Um, I didn’t get-I didn’t get that from any of the articles we read. (ID 15)
- “I would’ve loved to have seen more positive stories as time went on.” (ID 9)
- I need to know a little more about it. I need to know what the results of other patients, uh, are. And preferably other patients that fit my, um, same, uh, general condition, age, that kind of stuff, um, and whether I have to come down to, um, <Place> to have those done, or is it a one time thing, or over multiple days, or—I just need to know, you know, more general information. (ID 26)

#### *Hopes, expected benefits, and the existential threat of dementia driving willingness and readiness to start lecanemab*.

Stalls the progression

- Hopefully that it can stall this disease, the progression. (ID 5)
- it's not gonna—it—it's not gonna be the—the cure-all. But it could slow it down. And people could live longer, you know (ID 8)
- I'm hoping we get more life out of the people that think they're gonna pass out, you know, um, be—because of—because of this. Uh, or maybe they do pass away. But, uh, I'm thinking that if we could be a part of that and if we could, you know—Just slow it down. just, you know, just slow it down. (ID 8)
- That it may slow the progression, basically. (ID 10)
- the main thing is-is to stay in early onset as long as possible, is my understanding that that's—that it, you know, may help with that. (ID 12)
- It’s not probably gonna bring back any information for me, but it might help, um, you know, keep it from getting too much worse. (ID 3)
- **I mean, it doesn't cure it, so obviously I'm hoping that it significantly stalls it. (ID 5)**
- From everything I know, uh, eh, uh, lecanemab has a high probability—- of slowing decline (ID 9)
- I would be hoping that it would take the whole issue away.And I-and I know that there’s-that there’s no cure at this stage. But there—- there’s a slowing down. (ID 25)
- obviously **the hope that it can stall this disease**. **Um, at least I'd feel better if they could-they could give us some kind of parameters around. Okay, yeah. You know, for somebody with your subtype, you know, what we're seeing thus far, it can stall it two years, three years. I mean, if there were some kind of parameters around that, I'd feel even better. (ID 5)**

Help with day-to-day

- I can do day-to-day stuff. Um, but if-if I have to remember something, o—like, in one of the stories or—*[sigh]* you know, one of my tests that I took—with Dr. <Name>, they—she would read a story, and then I'd have to repeat the story to her. And I—that-that really frustrated me. I couldn't really do it. But when they asked me questions in-in—within the story, then I could answer that… if it helps, then that would be great. (ID 18)

**Does not want to be worse down the line like others**

- **No. I was gonna say, um, I mean, who—I mean, the people that I was talking about was just they-they can’t even talk. They can’t do anything. You know what I mean? So, that’s, you know—if I don’t—if I can get in the drug, then I probably would be a little bit better. (ID 17)**

Want to fight as long as possible

- “My [relative] died [in his 50’s] waiting for a heart, and I think that probably—as much as it sucked that he didn’t get one, I think the-the experience of that, you know, knowing that I’m the same age he is now, uh, he-he fought like hell, and it just didn’t work. So, I’ll fight like hell for as long as I can. And at some point, it's not gonna work, you know, *[laughter]* whether it’s early or late. And, um, and I’ll just, you know—I’m appreciative for each day I’ve got.” (ID 2)
- So I-I’ll be fine with that. I mean I’ll be *[unintelligible 23:26]*. I’ll say eff it, I’m gonna beat it, and even knowing I can’t. But, uh, you know, it-it—I-I don’t see myself getting depressed. But yes, the-the physical thing. And I haven’t-and I haven’t been overly stressed. I’ve gotten used to it. (ID 25)

Wants to be in a better situation

- Yeah, I am. I'm trying to digest that. Well, I want to be in a better sit—I really, really want to be in a better situation than I am. And if this has a good chance of doing it, let's get after it. Yeah. It's almost freeway all the way. It's more get on, get off. And there-there I am. If-if need be [someone can drive me], or maybe I can do it on my own. I don't know. Can you—would you suggest that I can do it on my own if I *[unintelligible 34:38]*? The first couple, and then what—then you get used to the drill and then I can go from there. (ID 16)

Biologically

- I've heard that the, um, some of the tangles—- are-are breaking down (ID 4)
- Uh, sure. Um, he basically—as I understand it, there is a study has come out that shows that this drug, when taken, uh, does reduce the amyloid buildup in about half the cases. And that-that amyloid buildup is what they think contributes significantly to the memory loss and other problems with Alzheimer’s. (ID 3)
- “I heard that the FDA was studying i- that the results were 25 to 30-plus percent, slowing of cognitive decline for, oh, 1 year or 18 months, period, and, , that they didn’t know beyond that” (ID 9)
- “the-the focus on the brain and-and remove the amyloid” (ID 11)
- Um, I don't know a lot about it. I just, uh—just know that it-it's, uh, an infusion— and I know it's every two weeks.It-it attacks the, um—the, uh—the globs that are in your brain cells and so forth. (ID 22)
- [I hope] that they’ll come in in a year and say, “We can’t find any amyloid plaque—- in your brain.” “And, uh, that, um, the-the tau tangles, or whatever they are, um, seem-seem not to be there.”And, uh, they, um, I, uh, they did not find tau tangles. They did find amyloid. And, um, you know, the—this seems to, uh, at least, ah, hold hope to reduce.And I would hope two years from now they’d come in and say, “Disappeared.” (ID 9)

It will get me back to where I was

- “That it's going to help me to get back to where I was, as far as—I don't really know the definition of mental acuity. **I have my own—that is how well you accept what comes at you. And mine has flipped. And sometimes what I think I'm understanding, I'm really not. But going over the same thing instead of once or twice—with the average person going over it five or ten times with me—I finally get it.** (ID 16)

More memory

- Yeah. I’ll get more memory. Not more, but, you know, like, little bit better. I think the doctor told me about that. (ID 17)

To help me and others

- Well, I-I think, to be honest, the number one thing, see if it'll help me. I also feel like it's something worth doing to see if it'll help others, too. You know, *[unintelligible 09:54]* working with things that come with this type of condition. (ID 22)
- No. I'm not worried about it.I don't think anybody would—nobody wants to put somebody in—in worse *[laughter]* condition. *[Laughter]* So—so I'm assume—I'm assuming that this—this would help. This would help me and many, many, many, many, many others. Um, and I'm sure that they're—that—I think that'd be the best thing. I—I mean, I wouldn't mind doin' that in a second (ID 8)

More time until there is more advancement down the line

- in the-in the interim, you know, like obviously frees up time for researchers to come up with even more advancements and, um, treatments and medications that can either further stall it or maybe help, um, stop it all together (ID 5)
- Uh, I think I just made the decision that, for me, um, not only because of the immediacy of the potential, uh, help from lecanemab, but I don't know what’s gonna happen in the future. And as long as I can, you know—hopefully, this gives me more time to see if something else happens as well. (ID 2)
- The other-the other thing that comes in is if you get on something, does it preclude the something better coming along?In the next two years you know, or whatever it is, that’s better. But because you’ve been on this or on this, you can’t do that. I don’t know how likely that is. (ID 25)

Independence

- I feel like I could manage going forward with the capabilities that I've retained thus far, um, to function fairly independently, um, if needed. So, I-I'm in a good spot, so if we can stop it in its tracks, that would be wonderful. (ID 5)
- I'm just hoping to have more time to be independent and, you know, enjoy life. I mean, we basically, you know, we live in a cabin. We're-we're on a lake, uh, you know, I have a boat and play some golf, you know, life is perfect. (ID 12)
- Yeah, I like to have independence also. I like to go shopping by myself, and we live in <State>, so we can go around to places, picking up friends and stuff. But, you know, I-I like my independence. (ID 20)
- . The truth is, I'm [in 70’s]-years-old and I-to me, my physical body is actually very good for my age, I think. But, I am greatly—not as good as I was 40 years ago**. I mean, I physically feel like I'm on a death bed and I'm not close to it. But you understand what I mean? I don't—I'm going skiing this next-this week-next week— -and I just have a feeling this will be the last time I go skiing**.(ID 10)
- Being able to do things independently. Um, you know, I know I'm not gonna be able to drive forever, but, uh, you know, I-I drive around town. I don't do anything, uh, you know, long distance, but, you know, I wanna be able to get in the car-in the car. (ID 12)
- I go down to the stream, do some fishing or go out in the lake fish or, uh, yeah, just-just the thing—you know, play golf and, you know, uh, I don't mind. It helps my golf game 'cause I forget a lot of shots. So, my scores have been but really good lately, but I also forget where I hit the ball. So, I hit the ball and I go, <Name>, where'd I hit that ball? So, she's gotta keep track of her ball and my ball, but yeah. Yeah, it's a-it's a comedy show sometimes out there.(ID 12)

Potentially going back to work

- Um, uh, potentially. I just-I just stopped working in December, so, um, and again, that—the way this disease has progressed for me, um, a lot, I was able to like camouflage it, um, because I worked at home. So, a lot of it, um, like with the disorganization and stuff wasn't apparent, uh, to people and I could still, uh, talk a pretty good game. (ID 5)

“To be able to live a life”

- , I would like to be able to live a life and be able to, um, speak and—and, um, not get wound up in—in my, um, my mind. My—my head. *[Laughter]* My everything.Anxiety. This is the anxiety. *[Laughter]* This is *[laughter]*— Um, so it's—it's—um, what I'm thinking it will do. I'm—I'm hoping it can help not only just, um, you know, not just the hope. But it's—it's good to have, you know, a plan and a—And a—and, "This is what I'm gonna do."And, "I've got good people around me." Uh, I feel good when that happens. Um—I have to say. (ID 8)

More time with family to do the things we love

- Um, I would hope to have a little-a-even a few extra time with the people that I love and everything. We-we’re-we’re try-we're gonna do some traveling. Um, you know, and—but it’s, um, you know, we can’t run out of money ei-either, too, but, um-but, um, we’re gonna try to do some trips. Um, that’s the big thing that we wanna do. ....just to help me stay be-better longer. (ID 4)
- **We're retired, and then bam, this hit, you know, and, uh, it-it's, uh, it's, you know, it's devastating. It really is. So, uh, the goal is to get as much good time and make use of it. I like to fish, play golf. We-we travel some, we're head to <Country> this weekend for our 10th trip in a row to <Country>, so those are the things that I enjoy. I** mean, I miss the drinking, but I don't need that. I still love a great IPA, but I figured it would just be easier just to quick, you know, altogether than to—I need the brain cell, the good brain cells that I still have. (ID 12)
- Just to do all of the—I don’t wanna say they were the bucket list kinds of things, but, you know, we just-just—uh, I think because my [relative] died so young anyway, I think we’ve always tried to appreciate all the moments we have, and we’re just trying to continue to do that. So. (ID 2)
- “I have a lot of hope that this will give me even more clarity and-and more, um, time with some clarity so that, you know, I can keep hiking and-and keep seeing *[crying]* my kids. Neither one of them have any interest in having children yet, so I don't know *[laughter]* if I’ll see grandkids. But my [loved one] getting ready to start her [graduate program]. My [loved one] is very busy in his job. (ID 2)
- To live longer. Uh, what else is there-is there? Well, I wanna-I wanna be there for my [loved one], um, my family, and, uh, ev-everybody that I, you know, love and all that. (ID 17)
- . I-I’m hoping that it will at least stop it in its tracks somewhat and, you know, slows it down so that I can enjoy more of my life before I don’t know who I am. (ID 6)
- Uh, I'd just like to, uh, I guess kinda get my brain functioning better and—you know, 'cause I-I'm not the same as I used to be at all. And so I'd just like to see if I could kinda get back to that level or similar to it or—I know I'm gettin' older, but, you know, just wanna function and be with my grandkids and family. (ID 22)
- “my feeling about it is-is that, um—and-and I’m anxious to get any feedback if I’m wrong on this—is that as long as I can tolerate it—and with the two APO4s, I think that’s still a question on the risk factors. But, um, if I can tolerate it, that, um, it buys me time, right? I don’t expect—this isn’t a cure or something that, uh, you know, it—I’m hoping to buy time, that maybe additional medication or therapies will happen to buy me even more time. Um, so, I’m not, um—I don’t think that I am, uh, pie in the sky, that this is a cure, or that this means that even though I’m only, you know, in my mid-50s that I’ll be around till I’m 112 or something. Um, but as much time as I can get is what I’m going for. So, um, that’s where I am with thinking that—again, I-I realize this is all still new, uh, these protocol-protocols and everything. (ID 2)
- Everything’s important to me. Yeah. Golf. Yeah. Or being able to, you know, uh—I’m never—I’m-I’m probably never gonna be, uh—I don’t think the drug is gonna make it—me perfect. You know what I mean? It’s not gonna—you know, I know-I know all that. So, yeah. I mean, that’s—you gotta deal with it. (ID 17)
- **I want more time, uh, wanna be able to get in the car and go put the waiters on, go fishing and I have to have somebody with me or, you know, wanna try to keep what I have for as long as possible** and, um, you know, you can kind of feel it progressing a little bit (ID 12)
- ’Cause my memory is getting worse. That’s the biggest answer. And if—and **I know there’s other people that, um, that we have and, uh, uh, uh—there’s a lot of people, uh, in <Location> have the same situation, and I-I know one of ’em can’t even talk anymore. I mean, ’cause they’re—they-they didn’t get it o-get it—get that drug. You know, too late, I guess. (ID 17)**

No hopes yet

- Well, um, uh, I guess I hadn’t got so far as to hope for anything. we’re-we’re still considering it. I mean it would be wonderful if it works, you know, if it really worked. But there-there just seems to be so many, um, obstacles or negative things going on. I’m just-I’m just not sure that it’s right for me. You know? And it’s the things we talked about earlier, you know, the—you know, where we live and autoimmune disorders. And, uh, reactions to drugs and things like that. (ID 15)

Going back to where I was

- To be mentally—to be really mentally back to where I was. (ID 17)

Is not aware of any memory issues

- Um, I-I don't know. Apparently, I might ha-have *[clears throat]*—excuse me—I might have some memory problems. I don't remember stuff, like I guess people do every day. I don't think that—think I have a problem.So I-I don't know what to say. (ID 18)

*Get better and not worse*

- That I will either get better or not worse. Um, or *[unintelligible 17:49]*—hopefully, *[unintelligible 17:50]* that would be the outcome. But if it would just even delay the process, that-that’s a win too. So— (ID 19)
- Well, I'm obviously—would be hoping for, uh, I guess, complete recovery from this Alzheimer's dementia stuff; but I understand that may not be possible, but we can at least make some improvements. (ID 24)

Recognizing family members

- Oh, recognizing my loved one would be good. (ID 3)
- Uh, well, not so much stuff I like to do. I like to be able to communicate with my loved one, and, uh, when she asks me, "Do you remember so and so," uh, and somebody I should remember—to be able to remember their name and stuff. And I-I do a lot, but, um, not as much as I should. Especially with , uh, the ones—people we haven't talked about or, you know, haven't thought about for quite a while. Um, those kinds of things, you know. (ID 26)

More time vs quality of time

- “He’s much more worried about, “Is this gonna actually shorten my time with you?” And I-I get that. I totally understand it. Uh, my feeling is-is if—and again, it’s because of the double gene, I think, that the risk, you know, is what’s making him—and-and I feel like, well, one step at a time on it, right? I mean, and to some extent, I mean, I suppose the very first time I tried it, maybe the worst thing happens. But, you know, we’ve talked a lot about it in terms of my feeling is if I can—if it works and if it—you know, we don’t know yet, right? So, maybe one and done. And maybe this is something that becomes a chronic problem, and it’s 20 years later, and I’m still havin’ a good time. I know none of us really know that, um, but we have a chance.” (ID 2)
- **It’s because, you know, I’ve had so much trouble with drug reactions in my life that, um, I’ve tried as best I can to stay off drugs and stay away from drugs. So I’m real—would be really concerned about, um, I’d rather have like two good years and-and, you know, with, you know, as clear a mind as I can have, rather-rather than having three years, but being so-so affected by the drugs that I’m taking that it wouldn’t be worth the extra year. I—does that make sense?**
- Okay. *[Laughs]*. Um, I-I-I guess I gotta leave it with, I'm-I'm aware there, you know, were some compromises and-and maybe, you know, further examination will reveal whether that's that's gonna do everything we want it to do, or I want it to do, have that quality of life plus treatment. So I guess that's not-not done yet, I guess is the-the bottom line. (ID 11)
- It-it also goes with a medication that was caused you to lose what you already have as far as your physicalness, your aliveness, those kinds of things. So if it’s a medication that’s going to maybe give you longer memory, but less life, that would make a difference. (ID 25)

Knowing it can do good for people

- That I've heard about it is something that could very well do—do good things for, you know, for—for people. Um, and I know that they're doing—maybe they're—maybe they're already started doing some and we just don't know about it, but it's—it's—uh, it's something that I think that a lotta people are hoping is going to be good. (ID 8)

Learning that it would not be good for people with autoimmune diseases

- Um, and the other one was that, um, the other kinda red flag was that it talked in there about, uh, that it wouldn’t be, um, good for people with autoimmune diseases. Um, I think they said active autoimmune diseases. And I have lupus, but it hasn’t been active for a number of years. So, um,but that still was a red flag for me. (ID 15)

Knowing it could bring memory back

- if it can help me, um, and get a little bit more memory—then that would be good for me. (ID 17)
- I guess, what I would—I said before is that my-my memory—um, I will get s-some memory back. But it’s—I’m not gonna get the most. I’m not gonna get everything. (ID 17)

Knowing that something can be done

- The fact that there’s something to be done or perhaps there’s something that we should consider doing, uh, because that was a different message than I had gotten from him the last time because the study hadn’t yet been released apparently.
- Well, that they showed, uh, I think the potential for helping people with Alzheimer's. (ID 26)

Latest and greatest

- “Well, we know that it's new and that we get-get—we're trying to get the approval and I guess that's all taken care of now. And it's supposed to be the latest and-and greatest for my situation, and we're just excited about, uh, going forward with it.” (ID 24)
- I think the biggest selling point was this is the best there is. Nothing any better and this is the way-way we have to go, or should go, and I was all for that. (ID 24)

Having a good attitude about it makes it better

- So—so our likelihood of—of, um, you know, it working for us we're—we're hoping is high. We've never had a, um, oh, and I think—I think we're hoping that attitude and being a part of it—makes us high in the likelihood of it working.(ID 8)

Statistics not helpful

- **Um, but-but again, you know, I’m not a, you know, I’m not a scientist or a doctor or anything. So it’s kinda hard for me sometimes to, uh, understand all the statistics they list. (ID 15)**

Statistics showed good outcome for half the people

- the fact that it did have a pretty good outcome for half the people that-that did go through the process. I think that’s a pretty significant improvement. (ID 3)

Doctor could have said more

- do something with the brain to help me remember stuff, I guess. Yeah. Basically. He probably could've told me more about it. (ID 18)
- Yes. I just could not think of anything how it's supposed to help my brain… they didn't probably tell me enough information about it. (ID 18)
- I would’ve liked to have heard, uh, more about the study earlier. (ID 9)

Nothing was not helpful

- No. Information can hardly ever be not helpful (ID 3)

Unsure how likely it is to help

- Um, I, you know, I'm not really, uh, sure about that. Um, I-I guess, uh , I would guess that that is-is, uh, is kind of still flexible at this point. (ID 12)
- “I’m not sure there’s any other—really, anything that’s shown that it-it could—and-and let’s be honest, I don’t-I don’t see this is a fantasy. I know it’s not a cure. Um, my hope is-is that it gives me more time and that maybe-maybe I don’t have—I’m not gonna be Pollyanna here again, but, you know, maybe something else comes along, right? And if I buy the time to-to get the crap off my brain and it helps me go along, you know, maybe something else will come along too.” (ID 2)

Has not heard enough about positives or negatives

- I have not. I don't-I don't know anything about it….positives or negatives, so I don't anything about it other than an infusion. (ID 18)

Brain bleeds

- the-the risk of brain bleeds (ID 5)
- I was talking about the-the-the bleeding, um, and the-my doctor, um, thinks it’s—oh, no, it’s not gonna be a big problem for me. (ID 4)
- Uh, yes. Specifically, uh, well, I would imagine there, um, the brain bleed is a big-big one. Uh, you know, who knows? You just, uh, you know, you react to the medicine or whatever, but it's, uh, it's a chance I'm willing to take. (ID 12)
- Or,you know, um,because the kind of side effects they talked about were things like hemorrhaging and, um, you know, other things that, uh, that happen. (ID 15)
- Blood b-bleeding (ID 17).
- think it's the-the, um, brain bleeds. Uh, we were—that was the one probably that concerned me or I thought about the most, but there's ways to treat that. And, uh—and I-I have all the confidence in the world that that's what they'll do then. So I think that was the biggest one that-that I was concerned with. But I'm-I'm confident that they'll take care of it. (ID 24)
- Um, well, obviously like the-the brain bleeds. Um, I know even though I'm not a carrier for APOE e4, um, I do know that obviously there are some cases where spontaneous brain bleeds, um, can happen. I'd probably want some more information on that. Um, from what I gather, um, brain bleeds, although serious, um, aren't that serious in the grand scheme of things. Like as long as they're small, um, you know, a lot of times people have them and don't even realize it. (ID 5)
- “And-and my first reaction was you gotta be kiddin’ me. But that’s because I-I went right away to the side effects.That-that was my first thing. ’Cause I-I’ve got other things going on and I’m always—that I’m takin’ meds for. And I’m always lookin’ at—- side effects” (ID 25)

APOE-4 gene

- "There’s a chance—I realize there’s a chance I won’t be able to handle it. Uh, the risk factor because of the-the double APO, you know, all of that will factor in. So, I don’t have some tra-la-la, you know, belief and a dream that, “Oh, this will just go great,” and-and, uh, you know—but it buys me time, and maybe something else happens, and maybe, you know—we don’t know yet how many times people could go through this protocol. So, one step at a time.” (ID 2)

Scans in place to identify brain bleeds

- and it sounds like the scans are in place to, you know, identify that [Brain Bleeds] (ID 5)
- said if one's detected, you know, thfey kind of back down the medication or, you know, and then if it's still a problem and they skip a dose and then, you know what I mean, there's di-sounds like there's different ways they combat that. And that even if you developed a brain bleed, it doesn't necessarily mean your treatment stops altogether. (ID 5)
- I did not, um—now without the tests, uh, at <Hospital>— - um, I might’ve, uh—if I heard there was a possibility of lung, bone—rather, brain bleeds—- um, you know, I’d-I’d—ee—oh—question things more. But for that percentage *[emphasizing]* and given that there’s a test that you can do that, uh, eh-eh-eh-eh—mu—demonstrates that you’re not, uh, you-you don’t have the, eh, genetic marker for—- that sorta thing—you know. I’m happy. (ID 9)

**Side effects in general**

- Um, he did-he did mention that there are some side effects, um, have occurred, some very bad side effects have occurred. Um, but that’s really about it. (ID 3)
- Well, I do know some of the-the, uh—yeah, some of the things that can occur. Um, and I'm-I'm prepared to go with that. They've—certainly they've got ways to treat those things. And I'm-I'm for—all for that. (ID 24)

**Death**

- “you know, strokes and death and all of that is possible. I am not thrilled about that” (ID 2)
- Yeah. They said that I could die. And I already—I kind of already knew that with, uh—I don’t know how I kn-know it, but my loved one, uh, re—she researches-researches all stuff, so it’s probably from her.(ID 17)
- I just know that-that there’s-there’s things that could go bad, and-and-and I’m-I’m ready for that. I’d rather-I’d rather be able to get onto the drug and-and-and-and whether—you know, I’d rather be able—on it. At least try it, and, you know, and if I-I die, I die. (ID 17)

Likeliness of side effects:

- Ten percent maybe. I’d-I’d still say 10 percent, maybe, or—- 5 percent or whatever. You know, I’m sure that the doctors are, you know, not putting you out there to die— (ID 17)
- “my opinion is that if that was, uh, clearer—clearly stated that it was 5 percent or 10 percent or something like that, um, as the number went up, I’d be more and more concerned. (ID 9)
- “it appears to be in the, you know, 5 to 10 percent range of, uh, of the database of people. Uh, something*—*it's-it's-it's something that'd be concerned with.” (ID 11)
- “I think that the brain bleed is a very low probability side effect, uh, because I haven’t heard to the contrary. And I think they would have mentioned that if, you know, if half the people got it, for example, I think I would have heard about that.” (ID 3)
- I think what he said it was a small percent. Yeah. It was a small *[audio cuts out* *19:21]*—some-somethin’ about proteins coming back.Yeah. It’s a test that they do to see if you’re more likely or less likely.(ID 6)
- That, uh, that-that he-he—it seemed to me he downplayed some of the, you know, the bleeding. Or something I saw that was-that was—- relatively insignificant.Or the likelihood that it would be—create harm was a-was a fairly low percentage or something like that. (ID 25)
- So brain bleeds are—Not unheard of.I-I-I think it’s—yes. I think it’s rare, rare even. (ID 25)
- I do know that obviously there are some cases where spontaneous brain bleeds, um, can happen. I'd probably want some more information on that. Um, from what I gather, um, brain bleeds, although serious, um, aren't that serious in the grand scheme of things. Like as long as they're small, um, you know, a lot of times people have them and don't even realize it. (ID 5)
- And the only-the other thing I had highlighted is, uh, look-does look like intracerebral hemorrhages greater than one centimeter in diameter have occurred in patients. And I guess I'd question like, what's-what makes it great-greater than one centimeter? **What makes that so bad? And like what-what to expect with that? … Um, so probably one of my questions would be, um, is that something I need to worry about or because of all the PET scans in the MRI scans I've had to date, is that not a significant worry? (ID 5)**
- That sounds pretty drastic. It could be. I'd wanna know more about why. I mean, on TV when they do commercials—- and they tell you all the side effects, you know, depression or death and all that stuff. So, yeah, I just would want to know more about it.Yeah. And if there can be blood in the brain, you know, is that—do I need an operation to get that fixed? Am I gonna die, or will that—is that normal? (ID 18)
- Sure. Um, obviously, I wanna know how prevalent the bad side effects are. Um I mean, if it’s a little discomfort on getting the infusion, um, that’s not a problem. It-it—the things that I’m concerned about are the things that would either, you know, make it far worse or sooner than letting it run its course. (ID 3)
- Um, I don’t know, um, what the possible side effects could be. He may have mentioned some of that, but I don’t recall it. Um, so those are kind of the-the things that-that he talked about. (ID 6)
- I'm not sure how you get the message across about, okay, here's the side effect and here's how it's gonna affect you or-or there's a range of things that, but I-I-I just think it kinda crafted, if you will, that uh, it's-it's not just a sentence or two. And so, anyway, that's a nice to have. I mean, it's not something that's overwhelmingly short*—*you know, an overwhelming shortfall. (ID 11)
- Um, but, uh, um, I think we're just pretty much at that stage where we just have to learn more. I mean, we don't know much about the study itself. We know kind of what the medication does, but the study itself so if we can 24:09 learn more, if there's things we need to do or-or things we can do to help it be successful and whatever. But we'll go off of what we can find out and what we learn. (ID 22)
- I'd like to look into it *[unintelligible 06:28]* more information, uh, you know, other than general information. Well, um, how have the test results, uh, been so far? Um, what are the risk factors, um, if any? Um, what-what's the profile of someone who is really a good candidate for-for the procedures? Uh, that kind of things. Improvement. So I mean, if you get improvement, uh, you want more improvement than less, but improvement is just what you want. (ID 26)
- Well, I'd want to know what the possible side effects are and how to deal with, uh—if I have to modify the medication, uh, or change it. Um, I mean, to modify, um, that's one thing, and to change it's a different thing. So to modify, um, I'd probably want to know if I can cut it in half or, you know *[unintelligible 08:27]* a half or, you know, uh, something like that. And to change it would be, uh, because I've already tried some other things with it and it probably didn't work. (ID 26)
- And what side effects are abnormal and I should stop taking? The normal ones I'd continue taking, but the abnormal ones I wouldn't. Right? You know, expect the doctor's input.Um, yeah, I—yeah, of course. Naturally. (ID 26)

Might make things worse

- That it's going to make me worse. Well, I want to—I want to do it. Unless you're telling me that the chances are just too great, that it's really going to—that it's really going to hinder me more than help me. But I don't hear that from you. I don't hear that from the doctor. (ID 16)
- I don't want to lose it. You know, I don't want to lose my memory and, you know, have, uh, backward movements. I think that's a concern. Um, I, you know—and I don't want it to affect my vision or, you know, other things, other function of my body. *[Unintelligible 15:19]* you know, I don't want that to happen without knowing the cause of its—you know, that it's causing a side effect. (ID 26)

It may interfere with life

- will it interfere with my life? Um, you know, that there’s some, uh, there’s some devastating, um, uh-uh side effect that, uh, hasn’t been discovered.

Uh, but, you know, e-ei-either I fall and break a bone and, uh, disintegrate from osteoporosis—- or I, uh, I go, uh-uh, from, um, Alzheimer's. (ID 9)

**Would partaking in this treatment stop me from any future treatments/medications?**

- And also—and maybe this isn’t even knowable. Uh, if doing this particular treatment would prohibit me doing other treatments that may come down, uh, over the next little period of time or bigger period of time that would be more effective. So, with one of these things is, do you get one shot at this? So, you could try one thing and if you don’t get a really bad result, maybe you can try another? And I don’t even know how you’d know that. (ID 3)
- The other-the other thing that comes in is if you get on something, does it preclude the something better coming along?In the next two years you know, or whatever it is, that’s better. But because you’ve been on this or on this, you can’t do that. If I start this-this one, does it impact something new coming along? (ID 25)

**Cost:**

- Well, I think Dr. <Name> had mentioned something about the cost, and we really haven’t heard any real details about that and how our insurance or—would—Would cover it or not. Yeah. Yeah. We--at some point, we wanna know the numbers, but that is not—unless it’s, like, a huge amount out of our pockets, that’s not gonna be a prob—the problem. Yeah. Yeah, I’d like to know—you’d like to know a little bit about it, but it-it’s not a main—a decision factor. (ID 3)
- the cost might be (a big problem for me) (ID 4)
- It's expensive.I-I didn't realize it could be that cost that early. (ID 11)
- Well, the biggest drawback is how much it's gonna cost. I don't know if I can afford it. My understanding was that, I mean, that there would be some, but I don't know what the price is and the cost— Out of pocket. That will make—that's-that's a stop right there, if it's too high**. I mean, the-the first thing and I really didn't think about that was the cost. That to me is the only thing that will possibly stop. And I think we'll know that when I get told how much it's gonna cost. (ID 10)**
- I’m not, um, totally aware of the cost of the drugs. Um, but you know, we would have enough money to do that if we wanted to. Um, but traveling, um, especially in winter here, is—I just—I don’t like to do that anymore (ID 15)
- My-my thing is if it—I don't have any money, so if it cost money, then I'd worry about that. And if I have to travel to <<City>> all the time, that would be a problem—well, not a problem, but inconvenient, probably. (ID 18)
- Well, definitely the-the, um, you know, the cost of everything. Um, I do have a good insurance.Um, they’re working on that now, as far as I know. Um, and, you know, so, you know, that’s always a concern. I know—I think he said it was like $30,000 every time you have a treatment. And so, you know, yeah, that’s a concern, but, you know, they’re workin’ on my insurance right now to, you know, be sure that all those things will be covered. (ID 6)
- Yeah, I-I wouldn’t want it to, um, deplete our-our finances. And I worry about that. (ID 20)

Infusion logistics

- it’s, you know, the wo-the worst negative thing would be that you gotta get there every week.And I think now they’re saying every two weeks.And, uh, you know, I have a life, and I wanna go—[loved one] and, uh, his loved one live in <city>.So the first thing I’d get when-when I got the, uh, I got the word of a date—- uh, for the first infusion was to—we’re goin’—we’re leaving on April 1st, uh, maybe the evening of, uh, the evening of Easter, and we’re gonna be back two weeks later. And I have my first infusion two days later. So—I-I’m—ah—I—if I had been able to get the infusion earlier—- we would’ve done something else. (ID 9)
- Sure. I mean, you know, if you just listen to what you just said, it's*—*you know, that*—*that's a big ask. And, uh, I-I-I-I don't mind saying it's something that, um, you know, decisions aren't final or-or maybe there's more that needs to be done, but there's a*—*there's certainly a-a quality of life issue and, uh, would like to think that there's something beyond what we're-we're discussing that allows us to still tr- still treat, but not, but not dominate or,-or remove entirely your availability for things to*—*you know, to do things that you really enjoy and like, so I don't know. So, yeah. (ID 11)
- “like cancer, and going in for chemo every day or every week or whatever*—*I think that's too often. (ID 23)
- it keeps you from doing stuff, going places, and things like that if you've gotta go to the hospital every two weeks (ID 23)
- And how much-how-how much hassle around the injections do I wanna have to go through? That’s a stressor, which isn’t my friend either.That, uh, yeah. And if we wanna go to Europe for a month or take a cruise for two months or somethin’, uh, that totally eliminates that. So.If there’s only one-third shot at it, I mean— (ID 25)
- Um, I think maybe knowing more about the infusion. Um, I’m not really sure how an infusion is done. Like, is it all just done through IV or—I’m not sure how that’s done. Uh, I would say from the people that are doin’ the infusion that, you know, know how it all works. Um, it—a website, and then if I had more questions for a doctor, I would do that. It’s a different team, right, that—- that does this? The—you have your-your doctor, and then you have the team that does the actual treatments. (ID 6)
- “I would say, and the recovery from, uh, you know, from that drug and its treatment, um, I-I assume there's a-a need for that. And I'm assuming also that there is*—*it is available and will be-will be part of my treatment. (ID 11)
- I mean, the frequency is something. Is it-is it gonna be every two weeks? Is it gonna be every two weeks forever? Or is it gonna be a shorter time period? Um, is-is—you know, that-that’s really—um, um, I can’t think of anything else. (ID 3)
- Um, I think it was helpful to know, you know, how often I had to go there ’cause we live a distance away from the hospital, which is fine. You know, we-we-we can manage that. (ID 6)
- But, um, and it’s my understanding that—I-I don’t know if he said we cannot skip one—- you know, for any reason. Um, and I’ll give you-I’ll give you a, um, an example. Um, my [relative] passed away quite a while ago now, about a year ago. Um, and I have not been able to inter her body down to, uh, <State>. And so my kids are gonna down there and do that, and I would like to do that. But I—my-my treatments, at this point, is my priority. So if they have to take it down—you know, take her down there and inter her, that’s fine. Um, but it’s just tryin’ to figure out some things that way. Um, and I know the treatments are every other week. So I don’t know. Is that gonna make me sick? Is it gonna—do we know if it’s gonna make me sick? Am I able to travel in between if I choose to do this with my kids? Um, they’re all adults now, so, um—so I guess those are some of the questions that I have. (ID 6)
- Well, uh, the traveling back and forth, it would depend on how much traveling back and forth. How often do we have to come down and come back home, go down and come back home, et cetera?They provide, um—if we need to be there, uh, instead of, you know, one week and then the next week—if we need to be there one day and then carry it over to the next day, does that provide, uh, compensation? *[Unintelligible 20:47]* and-and/or can we possibly get it here in <City>? (ID 26)
- I don't think so—- if it didn't hurt. Oh, I don't think it'd hurt my memory, but, uh, just the pain if it's an infusion twice a month, and—- you know, gettin' stuck with a needle, but—and I don't know how long that would take.So I guess I'm here to find out more about it then. (ID 18)
- **The only negative I would say is the—really the unknown, like, am I gonna get possibly sick from the treatment? Um, you know, do we know if I’m gonna get sick from—you know, like-like some people, like, when they have cancer, they know that they probably gonna get sick. Do we know, like, with this medication, if it’s gonna make me sick at all, or if it’s just gonna not affect me like that?** Um, is it going to make me sick? Could I die from havin’ a procedure like this done? (ID 6)

Stress associated with the disease, not necessarily the treatment decision making

- Maybe a tiny bit, but I think it's more so, um, stress associated in thinking about the disease in general...., you almost-you almost grieve, you know, what-what you potentially may miss out on. So—...obviously I'm a little scared, but yeah, no, I'm-I'm ready. Well, I'm hope-I mean, I'm hopeful that it obviously will stall it. Um, I know my doctor had said before when it—she was surprised that it took so long to get my initial appointment at <Hospital> (ID 5)
- I deal with stress every day. Do—like a lot. It’s just like, um—'cause I still work, and-um, I still work. I-I had to change some things for the-for the work, and, um, there’s a lot of people, um, that are-um, a-a lot of people are, um, uh, I re-s-s-um, they’re cheering me on, you know? (ID 4)
- Uh, oh, physically stressed. I-I have. Yeah, yeah. I mean if you call physically stressed wakin’ up at night and not goin’ back to sleep and thinking holy <explitive> .And I-I’ve had a lot, you know, with the hearing aids and the prostate cancer and, uh, oh, *[unintelligible 20:59]* the one thing I never wanted to get was a CPAP and that-that I got hit with this year. And just in the last few months so now I gotta blankety-blank CPAP machine. Which isn’t anywhere near as bad as I thought it was. But it’s like go ahead, give it to me, I don’t give a damn. I can take anything you wanna give me.And, uh, and I’m an emotional guy and I get emotion just sayin’ that. (ID 25)

Other medications

- So, um, it-it’s something you—actually, I think you should-should consider, but it-it—there’s no—I don’t-I don’t think there’s any precorn—you know, ahead of time decision of what—which one you would give up if you had to give up one. And if it is a-a-a symptom that could create a problem, is-is it different depending on which of the thinners you’re on? I know there’s two main types of blood thinners, and one is very well-tested and old and one new, but they both have different characteristics. And I don’t know. Maybe you’d have to pick one over the other because of the interaction you’re talking about. (ID 3)
- As far as I know, I don't think there are any major issues. Um, I was on some statins, but I'm off of them. So, I think, uh, I don't think there's anything that jumps right out, uh, that would affect me. (ID 12)
- Um, well, I take a baby aspirin every day. For, uh, one a day. You know, just a baby aspirin, one a—once a day for I’m not quite sure what. *[Laughter]* And because I don’t have a spleen and, um, have other autoimmune things, uh, they also recommend a baby aspirin for-for some of that stuff I think.But, um, anyway. Uh, and you know, everything I’m taking, um, now, the vitamins and the, um, the aspirin, um, not to-not to include the Memantine or whatever it’s called, uh, is optional, in a sense. (ID 15)
- ***Taking baby aspirin is “optional” (ID 15)***
- Coumadin, I take a little bit of it. Try to keep, uh, that under control. The doctor that I have, you know *[unintelligible 08:24]* they want to keep it so low. And it's like he looked at me one day and found another doctor that prescribes a high dosage of, um—forgetting the name of this stuff. I guess it is Coumadin, isn't it? And he kind of, you know, "I'm going to bring you back." It's a 40 mi-milligram, and he says, "Take three of them a week, not one of them a day." So he lowered that. (ID 16)
- No. Oh, I take aspirin, baby aspirin. Hmm. No. (ID 6)
- “Um, I take, uh, [dose] of aspirin every day, uh, but—and that’s for stroke.- uh, or the idea of-of possibly preventing stroke” (ID 9)

Stress associated with waiting for next steps

- I don’t—with everything that we’ve gone through with my [relative] and everything else that we had in our life, um, uh—I guess the-the answer is, yes, there’s been stress, and there’s been, you know, conversations, and there’s been times I’ve been really pissed off about the whole situation”

If I felt pain, could be stressful

- I suppose if I started doing it and I had tremendous pain, physical pain. I don't— I don't feel stressed....As long as something physically is not happening... I won't worry about it unless that happens. (ID 10)

**Tracking progress**

- The other thing that I would probably try and find out how—if there’s-there’s any way to do it is to see how quick the progress is going. Um, and I’ve only had one PET scan, um, and, uh, I don’t know if it makes sense to get another one down the road to say, “Well, it’s been—it’s-it’s staying steady or no, it’s getting worse.” And I think that would be a-a factor I would like to-to consider if it's-it’s able to—you’re able to do that (ID 15).

**Other treatment options**

- I’d like to know what other prog—uh, therapies are either underway in-in-in testing now or about to come out. Um, is there any, uh, refinements about the one that I’m going to be taking? Are they gonna maybe ha—do a study with a-a slightly different—instead of an infusion, a-a pill or just a regular shot every once in a while? Is that coming out? But those-those aren’t really big decisions. (ID 3)

**Nonpharmacologic interventions**

- “And also the other thing I’m curious about is other kinds of—I don’t—and I don’t even know, this may just be wishful thinking. If there’s other kinds of treatments that, um, I don’t necessarily mean, um, drug treatments, but **other kinds of things that you can do to help yourself. You know? Um, to help your brain function.** And, um, so that’s another thing I’ll be lookin’ into. Um, you know, tryin’ to see.**You know, and a lot of that’s kind of probably not scientific.** But, um, you know, but I-I’m thinkin’ about things like, um, medication, uh, to stop the—just to stop the anxiety. Um, not-not medication. Uh, meditation. Did I say that? **Meditation— to like stop the anxiety.” (ID 15)**

**Weighing risk**

Worth the risk

- “just in my own kind of weighing the pros and the cons, um, from what I do know and have heard, um, it's-it's worth the risk to stall this hopefully. Um, nothing else has been presented to me, so it's almost like, um, without it, I just kind of imagine like just my brain basically dementing at a faster clip.” (ID 5)
- Uh, what do I have to lose? Is basically the way I look at it. I mean, even if it does cause problems for me, I'm headed that way already. I'm not—might be a different kind of problem. I don't care. (ID 10)
- I don't care. Even if it speeds things up, I mean, that's a risk you take. I take a risk just walking down the street. And every one of us is at risk. That something will happen somewhere, sometime. So I don't worry about it. (ID 10)
- But, you know, I think some of it is because it’s so new and because it’s so, certainly, new to us, um, you know, we’re not gonna be stupid about it. I think it’s-it’s gonna be one step at a time. Particularly because I have the double gene, you know, um, I’m not gonna be really stupid about it. If it’s clear as we start it, that, you know, it’s not the right thing. We’ll have to make those decisions then. (ID 2)
- But, you know, my feeling was is I’ve had a great life. I’m only [50’s]. I’d like to go a whole lot more. But, um, but there’s a drug now, right? So, I think we both feel like—we realize that there is some greater risks and chances there. Um, and we may change as—you know, if something becomes more apparent that that’s causing a problem. But we just decided, what the hell? *[Laughter]* You know, I’d rather-rather see if it gives some help, and-and I still have lots of hiking to do. (ID 2)

“my opinion is-is that, um, at the moment, is that I’m worth the risk, even though I’m higher risk on it. So.you-you die at [50’s]. You can’t do anything about it when you don’t get a heart, right? Well, I can’t really—I can’t do anything about it. I can’t change this. So, I’m gonna—that’s why we go hiking every day and try to do everything we enjoy doing. And, um, you know, we’ll see. I think some of it will be as, you know—obviously, we don’t know exactly what the side effects will be for me, you know, specifically, and all those things.” (ID 2)

- I’d have to take it into account, uh, frequency, severity, that kind of thing. Um, but I might be inclined to take a higher risk on blood being too thick than losing my mind. (ID 3)
- No. I’m-I’m-I’m just trying to take this as just part of life and just an experience and we’ll go through it. And, you know? **Don’t worry about what you can’t change-- - ’cause otherwise you’ll, you know, ruin what time you do have. (ID 3)**
- **Yeah. I mean, you-you know, I try-I try to stay positive more—- than I-I look at the downside—- but I certainly know that I need to know what the risk, you know, to the benefit is, um, as far as that goes.** (ID 6)
- I mean, if-if you had a 50-50 chance of getting a stroke—- on the table on your first infusion—you know, I-I’d, uh, say, “We—maybe I should wait.” (ID 9)
- Um, I'd have to hear 'em [side effects] first, but, yeah, if it's gonna kill me, no, I don't wanna take it. - but, uh, you know, if it's, uh—if the side effects aren't too severe and so forth, I don't have any problem with it. (ID 22)
- So I would be looking at something that would have a significant impact if I’m gonna take the—take it. And-and by significant, I think the only thing that—my only expectation would be that it would-it would diminish the acceleration of it. (ID 25)
- So, um, you know, obviously cost is a factor, but, you know, my insurance obviously picks up the majority of it and, um, stalling this or-or, you know, uh, abolishing it ideally, um, is-is worth-worth the risk of the MRIs. (ID 5)
- I think I’m, you know, cognitively up with what people are saying to me and what-what-what I can read about or have my loved one read to me about. So, I’m still pretty confident that, you know—how do put this? I’m worth saving. (ID 22)

Fear of the alternative (not seeking treatment)

- So—and I'm-I'm young, I, you know-I have the early onset version, so I have, uh, you know, two [relatives] and, you know, things I wanna be around for. So—um, the alternative, uh, certainly scares me. (ID 5)
- And if-if-if it's being used for another drug and has side effects, you get the effects are, I guess. I think so.And, um, it's not something I look forward to, but it's better than having it. (ID 11)

Risk Analysis

- I'm ready to do it, willing to do it. Um, my family's, I think, understanding, uh, in—in that route—route—down that route.Um, is it a perfect world? Not always. Mm-hmm.But I'm fine with that. Calculated risk. *[Laughter] (ID 8)*
- I’ve certainly had many nights where I’ve cried in bed thinking, you know, this is really a big risk. *[Laughter]* Um, do my kids, do my [loved one]—would rather have me around, you know, longer and not take the risk? So, we’ve had those conversations as a family a lot, um, to say—and I think, ultimately, um, you know, I—if I felt like this was a-an absolute, you know, you’re gonna take it—and-and because of the APO4, I know I’m at higher risk. (ID 2)
- Um, my guess would be is it would be what—the same thing, probably, my [loved one] would be thinking ’cause I think we’re-we’re pretty much on the same s-space, uh, the same space about what would be a really stupid decision to make, right? I go in and it turns out that you have 52 brain bleeds, and you’ll—we all know you’re gonna die the first time. Well, who would do that, right (ID 2)
- Well, and-and that would get weighed off against what the benefit is and how certain it is. and what the percentages of those that it works with.So—yes. Could I interpret that that it slows it down by a third?The fact that it slows it by a third is fairly significant.So I presume that it means over time, it continues to slow it down.That was as far as it went in 18 months, I get that.But if you went 24 months, maybe it would’ve been more than that. Who knows?Uh, but the-but the one-third response is sorta counteracts that big time. (ID 25)

Would like to start treatment soon:

Sooner the better

- Uh, I'm ready. *[Unintelligible 43:27]*. Because I have [loved ones] and—and, uh, and I'd like to live as long as I can. (ID 8)
- I'm feeling the sooner, the better. I've—I, uh, really do believe that. I think, you know, the—the more you just sit there and do the—the, you know, "No, I don't wanna go that route." I'd rather just, "Let's get into it." You know, um, I would do it now if you told me to. Um, I'd—I'd be there.I'll find you. (ID 8)
- You know, so it—and-and I-I-and I want it to be-to be safe, so, like, you know, but that-that—but I want it as soon as I can. I'm pretty certain. You know, like, I feel like—I mean, I still have this disease, but I think that working to do something about it that would help (ID 4)
- It's—I've definitely felt like it progressed this winter. I don't know if that's just winter blahs or whatever, but, um, the clock's ticking (ID 12)
- if we’re gonna do it, um, I feel like the sooner we get going, the less damage is happening in my brain. So, if we can slow things down, you know, that’s just giving me more time to be salient and to-and to, um, enjoy watching my kids continue with their lives and everything else. (ID 2)
- Um, I’m 100 percent. I mean, I know there could be, you know, things that can go wrong, but I-I definitely wanna do it. (ID 17)
- Right away. I’m just-just ready. I mean, I-I-I-I don’t know how to say it. I just think if I can get on this drug, I’ll get-I’ll get a b—a little—uh, get a little better.That’s all. Uh, all I wanna do is get a little better. (ID 17)
- I’m ready. I’m just anxious to get started so that—’cause I feel like the longer it takes, I feel like I lose more cognitive abilities. And so I’m anxious to get started in the hopes that this will slow everything down.Because I feel like, you know, once you lose what you lose-you lose, you don’t get it back. (ID 6)
- Well, first of all, I'm pretty confident that-that-that I've made the right decision. Uh, my loved one and I both talked about it and I got her 1000 percent confidence level in it. We both agree, and I just—like I said, I just want to-to get better and, uh, make the improvements that I need to do. And, um, we're ready to go. We really are—and do whatever needs to be done. (ID 24)
- “I suppose I could wait later, but I also feel like, y-you know, the damage that is happening, I know at some point—I don’t wanna get to the point where I-I don’t have the choice anymore, right? (ID 2)

If recommended, im ready

- I don't know if there's a readiness for it or not. I'm ready. Probably 8, 9, or 10. If that's what they recommend, then—you know, so I'm with it. (ID 18)

I would pursue treatment, but I still have unanswered questions

- Yes. Would the caveat, I probably have a couple questions that I-I want answered. (ID 5)
- I’m not unsure. I’m-I’m-I’m leaning towards it. Um, but I do wanna get the information before I make any final decision. (ID 3)

Don’t know until you try

- Well, I guess there’s no way of really knowin’ until you try it. But, um, I don’t know. I-I just—like I said, I’m confident in the doctors at <Hospital>, and I-I just feel confident that if they feel it’s safe to do and safe to try, then I’m willing to do it. (ID 6)

Have not gotten to that point yet

- “You know, it’s just we haven’t got to that point yet of where we have to make that decision. And, um, we’ve been really busy with other things so I haven’t had time to worry about it yet. Or, um, you know, really think about it in depth, um, about whether to-to do that or not.” (ID 15)

Hesitant

- If I had to decide right now, I'd probably be hesitant because I don't know enough about the medication.So I'd probably have to say, "Get me more information, and we can go from there." But, yeah, I'd probably be hesitant. the over-dosage on the-on the medication that made me throw up so much—that would be—cause some hesitancy, but, um—but that was a—kind of a—I guess that would be—I'd say a medical mistake from our family doctor, where he put me on too much of it. (ID 22)

Does not think that the doctor would recommend it:

- So I mean, so I-I can be fairly clear on that. Unless-unless he sees the results and-and waivers from that, then I would very seriously consider it. But, uh, then-then-then other things would come into play about the-the side effects, all of those kinds of things. But right now, it-it doesn’t look to me like it was something that was gonna be recommended.

### *Individual traits and preferences, family factors, and degree of trust in expertise influence how patients balance risks and benefits.*

**Does not think about negatives**

- **I-I have the ability to pass over the negativity a bit. So my answer to that is-is I-I—when the negatives came up, I-I didn't listen very well…**I think I kind of passed over it. I was really centered on the idea that, uh, I was going to get better. I let the rest—if there was any rest, I let it fly by. **You know, being an [in 80’s] man that is cognitively challenged is-is not all bad. You're kind of a funny old bastard wandering through life. And, uh, I kind of like being that. It's not that bad. (ID 16)**
- I would say—- um, people are too scared of the negatives. I mean, i—u—I-I-I can’t—I haven’t heard anything, you know? It’s not going to, uh-uh, you know, ma-make me fall over dead. (ID 9)

Thinking positives, not worrying about negatives

- **I'm ready to go. Uh-uh, I mainly try—I'm trying to think of the positives that could come from it rather than, you know, what could go wrong**. I mean, it's, uh, you know, the writing's on the wall, so I might as well, you know, I'm not just gonna sit back and accept all this if something that can be done. (ID 12)
- I just know that they were—remember saying something about that, but that fell in another category, I don't care. Even if it speeds things up, I mean, that's a risk you take. I take a risk just walking down the street. And every one of us is at risk. That something will happen somewhere, sometime. So I don't worry about it. (ID 10)
- Uh, it-it wasn't even a—it was like an instant. Yes. It wasn't even—I didn't even consider any-any of the negative things. I just, uh, you know, I—it's not gonna get better just doing what we're doing now. So, we as well try something. Yeah, it's just—I dunno if you heard that, but yeah, the anxiety is, when-when do we get it? Are we gonna get it in time? The clock's ticking, you know that that's (ID 10)
- **I-I have the ability to pass over the negativity a bit. So my answer to that is-is I-I—when the negatives came up, I-I didn't listen very well…**I think I kind of passed over it. I was really centered on the idea that, uh, I was going to get better. I let the rest—if there was any rest, I let it fly by. **You know, being an [in 80’s] year-old man that is cognitively challenged is-is not all bad. You're kind of a funny old bastard wandering through life. And, uh, I kind of like being that. It's not that bad. (ID 16)**
- I really don’t have concerns. I know-I-I know if—uh, you know, I can die. **And if I d-die, I die. (ID 17)**
- Um, you know, I don't really have the concerns. I-I-I'm looking forward to improvement, to get this thing taken care of, and I've got all my trust in what I've heard and the-the care I've already received. (ID 24)

**Is a “negative person”**

- Well, since I’m a negative person, sometimes when I’m looking at things, I know it was-it was only good in half of the-the people. Um, I don’t know whether there was any correlation, or any study given to, uh, how far along the people who had the bad result were, uh, or if it’s it doesn’t really matter. Whenever you stop the therapy, you either get the benefit or you won’t. (ID 3)

Trust in the doctors/professionals

- , I trust my doctor and the staff, um, and if something grows ba-goes bad, um, you know, I’ll have to deal with it, (ID 4)
- **You know, I, I feel, uh, I am gotta think the FDA has done its job. Like I think that you guys are, you know, have one of the-the best hospitals** in the world and the best doctors. So, I-I—as far as if you're gonna have this disease, I think I-I have it at the right place with the right hospital, with the right doctors, with the right partner, so, uh.(ID 12)
- Well, the one is an emergency room doc.And they're kind of cavalier in a different sort of way anyway. And his idea is-is that he leaves it strictly up to me. And I think the bottom line is, "<Name>, you're going to be all right. Uh, you're going to get by." And I say to him, "Yeah, with your help, with you leading me. Literal—almost literally by the hand." "No, no, no, no. It's not going to be that bad." "Well, how the hell do you know? You're an emergency room doctor." "Yeah, but I can—I-I have all this inform—" He's <Organization>, <City>. "And, uh, I have all of this stuff. I've had all of these years of going through it. What you're getting from me is not just anybody." Which I suppose you would agree with that, wouldn't you? (ID 16)
- I-I’ve always trusted my doctors. You know, and-and I don’t think they’re gonna, you know, go do something wrong or anything. So-so, uh, uh, it’s—I mean, I don’t know how to answer that. **I don’t have any fears. (ID 17)**
- Again, one of my policies is, don’t worry about what you can’t change right now—- as best you can. Don’t worry about it. And when we talk to Dr. <Name>, I guess we’ll know more, and maybe I’ll have some concerns then that I don’t have now. But I’m not gonna try and make up a concern to have to worry about for until I get to see the doctor in a month. So—Trying to live day-to-day. (ID 3)
- I've, uh-I've read that on my own, but it's also might be because I'm not that far along in the process. I do have an appointment this upcoming Monday with the nurse practitioner at <Hospital> to sign forms. So, I assume they would, um, would educate me a little bit more on it as well. (ID 5)
- I-I would think that, uh, before I start taking anything, I would, um, either get ahold of the doctor or do a little research. Um, you know, I don't wanna do anything that's gonna counteract the benefits of the lecanemab. (ID 12)
- I don’t know that. No. I-I-I-I, for me, I trust my doctors.Well, I don’t think I’ve had any issues. Uh, I’m not su—I mean, my loved one takes care of all the-all the medi-medicine. So—I’m not—she says I’m not o-on anything. (ID 17)
- I’m not really—I-I haven’t given that much thought. Um, my—the usual way I-I try, and approach big medical decisions is to do as much research as I can, listen to what the doctor says, and at the end of the day, put it in what the doctor recommends ’cause, um, they’re better at it than I am. They know more.	(ID 3)
- Um, and then with my doctor's agreement, I mean, given that the doctor said I was a good candidate, you know, obviously I would've hoped that, um, some time and consideration was spent on their end (ID 5)
- I know that no doctor is perfect. I **do honestly believe that they know a lot more about this than I do— -in many ways. And I trust that they're very, very good.** And that's the only way I know how to answer....I don't even think about it because I trust it. That's what I know. Unless something went wrong, then it might change. But **it's like, I trust the doctors. I really do. And there's no reason for me to not trust it.** It's like I say, I feel like something happens that makes me lose that trust. That's hard to get back. (ID 10)
- The doctor at <Organization>—finally, I work him around a bit. I'm talking to a [medical professional], and I'm saying I work him around a little bit. Oh, yeah, right. And I finally got it out of him. Well, why in the hell wouldn't you do it? Those maybe weren't his exact words, but they just as well have been. Got it. So the doctor kind of gave-gave pretty convincing kind of feedback into, you know, this*…*Yeah. Yeah. Well, you-you can't do any better than that, can you? (ID 16)
- Well, I'm fully aware that there could be negatives, but so far I'm willing to set that aside. I just want to do it. I'm leaning somewhat on my doctor. I think he said it all when he said, "Why wouldn't you?" That's all it took. Which, if you want to look into that—that's actually, on my part, pretty good trust in him, isn't it? (ID 16)

Physician encounters- pressure

- “[Pressure is] Just me. *[Laughter]* No. I mean, I think—I would say, actually, the, uh, the doctors have been wonderful. They-they have really explained the-the risks and the potential benefits. Um, if anything, I would say they were, um—I don’t wanna say that they were negative about it, but they were really trying to make sure that we understood the risks, um, of taking the med—particularly because of my double gene. Um, I-I won’t say they were trying to talk me out of it, but I—lots of conversations have happened to make sure that we all agree that we understand what the risks are, what could happen, what could happen, you know, in terms of lying there in a coma, you know, all of those sorts of things.” (ID 2)
- “I would suggest that my doctor probably, um, thinks I may be taking a little more risk than I should. *[Laughter]* Um, but-but it's been, uh, it’s been a very open conversation. And-and I feel like that I really understand, um, what I’m getting into, and I feel like that she understands that I really am okay making the decision.” (ID 2)
- Um, well, the doctor in <City> who I, you know, I really like— but he-he—I kinda got the impression that he was recommending it or that it would give me more years. Um, or that pushing it’s too strong a word, but that he was encouraging it in some way… I got the-the-the impression he was definitely pro doing the drugs. Um, and **I don’t know-I don’t know if that’s based on good results he’s seen in patients or-or whether it’s just kinda, you know, the latest thing to try. The latest thing to do. You know?** I don’t know. I don’t know yet. (ID 15)

No pressure

- You know, not really. And that's very reassuring for me. My friends, while they have an opinion, they're not forcing it on me. (ID 16)
- Well, maybe if it's pressure, just somethin' to do, to take it, but I don't feel any pressure. I mean, if it'll help, then that's not a pressure, but—Yeah. I mean, I don't care if I take it or not, but if it's gonna help, that's—maybe it's a pressure that I should take it or should do it. (ID 18)
- I mean, she doesn't say, "Oh, you'd better take it. Uh—" You know? It's not like that. So if it's somethin' I need, then she would back me to take it or—I guess. But I don't know what you mean by pressure. Nobody's pressurin' me to do anything. (ID 18)
- No. Not at all. I’m the kind of person that I am very adamant about my own decisions. So—like, if my [loved one] were to say one thing, I might say another, and that—if-if that’s the way it’s—you know, he might not like it, or I might not like it, but it’s my decision—- so yeah, very much so. (ID 6)

Pressure that there is something out there now

- “my oldest son when we were together at a-at a-at a place and I mentioned that to him. And he-and he—and this is the guy with training in this.And he said, “Well, [family member], what-what are you doing about that?” He said, “That’s because-because this is, you know, progress—there’s things that can be done that can slow it down.”And so that’s-that’s where the-the pressure thing triggered it” (ID 25)

Cost/Logistics

- I guess the combination of my, um, checkered medical history *[laughter]* and of all the, you know, the things I’ve been through. Um, and not wanting to do, you know, try something else that may or may not be-may or may not be helpful to me. Um, and also, uh, you know, just the effort it would take on my [loved one] part ’cause he’s the one who does the car… But just the, uh, the distance and the, um, the, uh, the-the hassle, I guess you’d call it, of actually goin’ through that process. I mean if it were down the street it might be one thing, but it's not. (ID 15)
- I think, um, you know, we both had good jobs. We don't have [loved ones]. I mean, we're not rich by any means, but we're, you know, we're comfortable. Um, so it-it—that's not really a-an issue at this point. Um—We're supposed to have insurance coverage. And-and, uh, yeah, our—it looks like, uh, our insurance is gonna cover it. Uh, so, you know, it's just a matter of the expense going down there and back. And-and I imagine once we're getting the infusions, I-I, like I said, the first time, we will probably stay down there just 'cause we don't know how I'm gonna react, but it-it-it's just a nice day trip. Um, <Name>'s best friend lives in <City>, so we, uh, we hook up with her while we're down there and usually get a meal and, you know—Explore city. (ID 12)
- Uh, sounded like the medication alone was like $26,000/year. Um, not factoring in like obviously the, um, hospital time and facilities and their staffing that's needed too. Um, administer this, obviously there's a cost associated with that. So, I mean, if it was out of pocket it would be obviously a problem (ID 5)

Previous experience within the healthcare system

- Also, my [family member], uh, passed away from, um—can't even think of the name of it now. Uh, begins with an A, aneurysm. Uh, so obviously when they did all her scans, they did say, you know, they could see some that were on her brain anyways that had, um, just I guess been dormant or whatever, were not a problem. Um, so probably one of my questions would be, um, is that something I need to worry about or because of all the PET scans in the MRI scans I've had to date, is that not a significant worry? (ID 5)
- “I don’t generally use very much medication anyway. Um, and I would say, you know, the occasional time I’ve had a headache and took some Advil or something, I could lay down with a-a w-wet wash cloth or something and be okay too” (ID 2)
- I think, probably, because, you know, my [relative] dying [in his 50’s]waiting for his heart, we had to deal with all of those kinds of issues and thoughts a long time ago. So, it’s—this isn’t the first time we’ve-we’ve had these kinds of issues. So, I don't know if that makes it—I think it’s easier for me, honestly, than my loved one because I went through that. (ID 2)
- Um, I missed my sophomore year in high school because I had, um, a [medical issue]. And they eventually did surgery and removed that and then I was fine. And then a few years later, I developed lupus. And I think those are both considered autoimmune conditions. In the meantime, uh, going through all these years of being sick and being in and out of the hospital so much, I had really adverse reactions to a lot of drugs. Um, to the point where today I don’t wanna take anything Um, because I-I’m so worried about side effects and wind-windingup back in the hospital, um, from a side effect. So, um, that’s amajor, major concern for me (ID 15)
- Yes. Yes. I mean, it-it—I think it [family members’ Alzhemer’s] has influenced every real significant medical condi—you know, decision. There haven’t been a lot. So, I-I’ve been—you know, knock on wood. I’ve been pretty-pretty lucky on the—in that front. (ID 3)
- “I have had, uh, relatives who died from, uh, brain bleeds—- uh, given the wrong medicine. Uh, but the-the numbers are off the chart. You’ve got a—you’ve got a, uh, high probability or might—de—hmm—a high, uh, likelihood, um, that you’re going to, uh, i—uh, at least, uh, slow this thing down. And at most I’m planning on being in the study or, uh, whatever—- uh, for anything that—- will slow it down or even reverse it. (ID 9)
- they-they put me on some medica—uh, kidney medication for my—to help me with my thinking, and it caused me to throw up all the time. Uh, yeah, if I don't—the only one I can add is the medicine that made me throw up. That was miserable, but besides that. (ID 22)
- “She[sibling] was very, very bedridden. Uh, they finally had to put her in a home. And, uh, she was the last sibling that I had was still alive, and-and she meant the world to me. And, um, she left a family of—and her-her children are now kind of our adopted children. You know what I mean? They're all a bit older, but, uh, it's-it's just left a big hole in the whole family. And I don't want to be—do that on this side, you know? I want to be here for everybody as we get older.” (ID 24)
- My [relative] lived to [his 90’s].And, uh, he, um, he, uh, fell off a chair and, uh, and just, you know, um—they wouldn’t operate.And I—m—uh, and he died.And, uh, um, I’m competitive.And I wanna beat him. (ID 9)

Role of Family

Alleviating any concerns of loved ones

- **how I rationalize that in my mind and then turn around and had a conversation with my loved one, I said, "Um, if I had breast cancer, um, nobody would question if I was going for chemotherapy tomorrow, but technically chemotherapy is a poison." And at some point long before we became conditioned to hearing about chemotherapy, it sounded big, bad, and scary too. (ID 5)**
- I mean my family has obviously voiced their concerns, but, um, once I kind of laid that scenario out about the chemotherapy, um, I think it kind of put everything into perspective and nobody's really said anything negative since, or, you know, said, really, we don't want you to do this or anything like that (ID 5)
- my loved one has been wonderful about it. I think he has more worries about the risks than I do, um, partly because he’s like, “I’d rather have you even if you don’t know who I am anymore. I’d rather have you as long as I can.”
- he says, “If this is something that *[crying]* you really wanna do and potentially it’s”—I don’t think either of us realistically says, “Oh, this gives us time till something else happens.” (ID 2)
- I suspect if you asked him, uh, and I wasn’t in the room, he’s probably more worried about it than I am. Um, but we’ve spent a lot of time talking about it, you know, and me included saying, “Uh, uh, if you just can’t deal with it, we won’t do it.” Um, but until—you know, we still have an MRI to go and a lot of other things. And we’ll see what the risk really seems like right before we start it. (ID 2)
- Mm-hmm. Well, um, so far it’s just kinda been my loved one and I. Um, you know, dealin’ with it and talkin’ about it. We haven’t really talked to a lot of other people about it yet. I’m just kinda waiting to see what, you know, what a, you know, what a—I don’t know if you call it a diagnosis or prognosis or whatever (ID 15)
- I, you know, really very good. Excellent. She’s sitting next to me. No. She’s doing a great job. I’d be lost without it. (ID 3)
- Um, they’re all very positive about it. You know, they-they feel, you know, positive, you know, that this is a good thing for me to-to do—- um, and a hopeful thing. So they’re all really, you know, supportive all the way through. **They can’t wait for me to start. (ID 6)**
- Um, uh, well, I’d said I-I’m concerned about my declining, uh, smarts or whatever, um, to my loved one. And she said, um, you know, “I don’t really see it,” but she, uh-uh, but-but I did see her, uh, concerned. And when I went to the—well, so I made the decision for me and—but-but she’s been supportive. I’m—very supportive. Um, well, as for my loved one, she said, “You know, this is a big deal. I’d like to go to the appointment.” And I said, “That’s why I ask,” and, uh-uh, but I mean, I think that once I get into this, um, I’m gonna try to be s—y—uh, point out to people, uh, that there is hope. And, uh, so I-I’m gonna be a billboard. (ID 9)
- Well, my, uh—my loved one has been the best support. And she's, uh, the biggest reason why we're going through a lot of what—you know, we're trying to fix things and so forth. Um, my loved ones. I have [some loved ones] in the area who are concerned about me, and a son-in-law. (ID 22)
- Well, our children. They all live—two of them live here and one lives in <state>, and the <state> one calls every day. And the kids here we see several times a week, and they're all supportive. Um, all my—all my, uh, [siblings] are gone. But we have <name> family, who mostly are around the <city> area, and we talk to them mostly every day, and they're all supportive. (ID 24)

Family understanding of my situation

- Um, I think it's just, uh, a kind of more of an understanding. Um, when I—there are moments when I'm fully fluent and I can do all sortsa things, and then something strange happens, and I—my brain says, "Oh, I'm gonna go for a nap.(ID 8)
- my loved one agrees that we wanna do it. (ID 8)
- Um, you know, we have a distant family here year-round and, uh, close family here most of the summer. So, we get support in that-that way. But, uh, you know, it's, uh—our life would be perfect if it wasn't for this. And so, if life sucked and then this happened, that would be really bad. (ID 12)
- Well, my [loved one], obviously. No. I mean, I-I-I couldn’t go any—I-I couldn’t do anything without her.No. She—I mean, no. She’s—you know, she takes care of everything. Takes care of me. Takes—does everything, you know. Sometimes I don’t need to—I mean, I can golf myself. We talk about it all the time. I mean, when—especially when we were going to <Hospital> all the time.(ID 17)
- she refocuses my attention. I'm spending too much time doing something, she'll smack me up alongside the head. Not really. *[Laughter]* Correct my attention and, you know—yeah. So that kind of stuff. She is wonderful. (ID 26)

Family matters most in this decision

- “My family.” (ID 8)
- Okay. Um, I—a big thing is my children. Um, they’re [teenagers] and I still wanna be around for them. And, you know, the, um, I want to, you know, continue-co-continue to, um, you know, be with my family and-as long as I can. Have, you know, good trips or, um—and I'm-I still like my work and everything. Um, and so, yeah, so-you know, and I-and I have a-al-also, I—for two years, I walked in the Alzheimer’s walk—and so, the first year we raised $10,000. I-I like to make a-a difference. (ID 4)
- “I’ll get to see my kids more often, but—um, is we’ll just try it, and we’ll see how far we get-we get. And, um, you know, we still have a whole lot of hiking and lot of fun things we wanna do. And, uh, if you asked my [relative], I would say he’s much more worried. He is, um, you know, “I’d rather have you forever.” And while, uh, that’s wonderful to hear, um, I’d rather have as good many years as I can have and have a shorter life *[laughter]* than just be in a bed and not even know who I am.” (ID 2)
- Well, I'm learning to live with-with the fact that I'm not who I was, and it isn't all bad—as long as my loved one is—I mean, she's not thrilled that, uh, I'm not who I was entirely. And, uh—but I think she's getting used to it. (ID 16)
- **I'm still there. I'm still her buddy.** But, uh, coming from a somewhat different angle. She's younger than me. So I think she's kind of looking at it from the standpoint that, uh, I'm using it as an excuse. And, uh, you know, the reality of it is, is I'm-I'm not. Uh, it's for real. (ID 16)
- Um, really, I think what matters to me is-is my family. You know, um, we’ve had lots of talks about things and, you know, um, you know, helpin’—I’m tryin’ to understand everything myself, but, um, I want them to also understand, you know, the risk to benefit and, you know, what the potential future could be depending on what happens, yeah. And we’ve talked a lot about that stuff. (ID 6)
- Um, I'd just like to be able to do more to help my loved one and to help my family and be able to function a little more. Like I used to like to sing a lot, and now it's kinda not there, and things like that. You know, get some of the old good habits back that I used to have. (ID 22)

Family members in the medical field

- Um, my sibling’s a [medical professional] in, uh, <city>. She-she's actually the head nurse, so she, uh, she's been a resource as—especially, I think <Name> has relied on her a lot as a resource. Well, I think, um, [sibling]s been a-a good resource I think for <Name>, right? Oh, especially in the beginning. She really helped me get going with the navigation, and then I just kind of took the ball and ran with it. Yeah, I mean, she's—she was on the call when-when, uh, I got the diagnosis. Uh, but, uh, <Name> had al-already knew. I didn't know, she knew. (ID 12)
- Um, really just havin’ the confidence in-in my doctors and, you know, um, I have a [relative] that works there in [medical department]. And, you know, she’s—you know, she-she talks to me a lot. She’s-she’s not in that department, but—- um, but, you know, she knows a lot. And, you know, she’s confident in what she’s—what she understands is happenin’, so that helps me understand.Well, she an—you know, she-she’s supportive—- of—- you know—like, she may not know a lot about—she works in [medical department]. But she may not know a lot about your study, but she knows, you know—she’s comfortable, and she-she feels that I’m in the right-right place. (ID 6)
- Our [family member] is… a [medical professional] he runs the tests and all this, exactly this, but with younger people.And he’s been very helpful with phone calls and his network that he’s got fairly wide, going to conferences and things, talking with people. And has been instrumental (ID 25)

Does not want to be burden on family members

- Well, like, if somebody's talking to me and I need to relay the story or an experience, I'll get the details wrong. And if she's around, then she'll correct me and help me out. So if she thinks I—if this would be beneficial to me to solve all that stuff, then that would be great. So she—yeah, she's there with me all the time.
- I would rather be the one with the burden. *[Crosstalk 33:18]* yeah. Yeah, that’s—that doesn’t mean—I-I’d rather have you have it, then I could take care of you. But, uh, I think knowing that you’re the burden—I suppose what the good thing is *[unintelligible 33:33]* get far enough down Alzheimer’s so you don’t realize it. I don’t—I’ve seen people like that and known of people like that.Don’t recognize anybody anymore so they probably don’t realize they’re a burden. (ID 25)

Friends as a support system- helping them understand this process

- They’re r—I mean, all my friends are really good. And my-and my family is great. Oh, yeah. I mean, like, **somebody, maybe, asked me, uh, “Yeah. What it—what-what happens if, uh, you know, something goes wrong?” And I said, “Hey. Tha-that’s the way it is.”**We’re all gonna die sometime. (ID 17)
- Um, a lot of my friends will pick me up, uh, and, if we’re going somewhere, and—you know, if we’re going to a watering place to go to have a beer—- and, you know, I-I got so many friends that would do that for me.Yeah. And I-I told you, I got too many friends. Not kidding. *[Chuckles]* I’m not-I’m not saying I’m—everybody loves me, but I do have a lot of friends, and they’re still friends from long ago. (ID 17)

Religious support

- “So as far as I feel—and then our—in our church, we have wards that we live in 18:11, and the ward members have been really good support, too. So I have a good, solid base for support.” (ID 22)

Decision Making

My decision

- Well, I kind of think I’m in the driver’s seat. You know? No one’s gonna force me to do it. Um, and I think it’s certainly better than not having any decision at all to make. So no, I don’t think there’s any stress or any problem around that. (ID 3)

My doctor and family will help, but its ultimately my decision

- **Was I will decide on my own— -with agreement from the doctor—is that one of the— I-I'm certainly willing to listen to any concerns from my family member, but ultimately it is my body, my decision**. (ID 5)
- I think it's a combination of two, uh, two things. I'd like to talk to the doctor—to understand what he or she knows and take that information with the family and say—listen, there's no—there's no guarantees. There's no guarantees. But I honestly think that, uh, my family would say, "Go for it. Go for it. Got nothin' to lose, really, in a sense (ID 8)
- I think overall, she's very supportive. We haven't really talked about it. I mean she's the same way. I mean in the sense that we don't know what is, so I don't really—can-cannot answer for her. My loved one doesn't even know. I mean, she may**—I would be glad to hear from her but I'm gonna be the one that decides. (ID 10)**
- I've decided on my own and I hope all those other, uh, shareholders or stakeholders, uh, agree with my decision. (ID 12)
- I’ve had enough experience with-with doctors to know not to-not to necessarily trust their opinion, you know. I’ve had some bad advice from-from doctors. All I’ve been through in my life. And, uh, you know, so I definitely would not just, you know, blindly rely on one doctor’s opinion or whatever. (ID 15)
- Oh, me and my loved one.Always that way. Yeah. Yeah. I—we talked about it and said yeah. Both of us. Yeah. Just the two of us. (ID 17)
- Okay. I would want to hear from Dr. <Name> and get his professional opinion. And then, uh, anybody else, you know, wouldn't be professional, so I-I would like to go off of what he says, and then me and my loved one or somethin' decide if that's what I really want or not, or if it is necessary at this point in my life. (ID 18)
- Obviously, what my loved one thinks is very important to me ’cause, you know, this is gonna affect her maybe not as much as me, but it will certainly affect her. So, I care about that. I care about what the doctors tell me because they know more than I will ever know. Um, but if I get a bad vibe or bad feeling about whatever is going on, it’s gonna be my decision. I know the doctor makes you say that ’cause it—he doesn’t want that accountability for it. But I really mean it. I think I will be one that makes the final decision, but I will take—what my loved one thinks is very important and what my doctor thinks is very important. (ID 3)
- I think we’ll decide after listening to our doctor’s opinion. (ID 20)
- I think us, along with the doctors, is how we would want it to function. (ID 24)
- I would say that, um, uh, um, we're—uh, my loved one and I are both in agreement, and as long as Dr. <Name>, after reading all this information, feels it's appropriate, then we would move forward. (ID 26)

Family and I will make the decision

- “I would say most likely it will be a we, me and my [relative] making the decision. Because if he ultimately—if some other, you know, oh, by the MRI or something, you know, it really is much more, you know, I will take that into consideration. But I would say, uh, overall, I think I’ll probably have the final decision.” (ID 2)
- It’s—- mostly my family. Well, and me. Me. I'm the— - most important person.” (ID 4)
- Um, as long as I got to “yes,” it doesn’t matter. Um, I-I think, um, I ge—it—having—bottom line, I made it with my loved one.But I—eh—m—but—m—would have made it on my own if need be. (ID 9)
- Yeah. I-I think it'd be me making it with my spouse and one other family member.Could be*—*actually a couple more 'cause uh, my [loved one] is now of age that we feel that it's good to get him in for his own growth development, but also to*—*he's-he's-he's the one that can, uh, fill in a*—*the empty slot that this creates. So, um, so something that says, I will make it along with a two, three family and-and, uh, family that would be with us. (ID 11)

Quantitative

MGH: 2-12, 17 (12)

Utah: 15-16, 18-21, 23-26 (10)

Ready to make a decision: 8,4,10, 12, 16, 17, 6, 9, 24, 7, 19, 26 (12/22)

Ready to make a decision but has more questions: 5, 2, 18, 3, 22 (5/22)

Not ready to make a decision: 15, 20, 11, 23, 25 (5/22)

Yes to treatment: 5,8,4, 10, 12,2, 16, 17, 3, 6, 9, 24, 26, 7, 19 (15/22)

No to treatment: 15, 23, 25 (3/22)

Not sure, need more info: 18, 20, 11, 22 (4/22)

Friends/family with provider: 15, 16, 12, 25, 6, 12
